# The COVID-19 pandemic and child malnutrition in sub-Saharan Africa: A scoping review

**DOI:** 10.1101/2021.07.21.21260929

**Authors:** Paolo Sestito, Sabina Rodriguez Velásquez, Erol Orel, Olivia Keiser

## Abstract

**Background:** Although the COVID-19 pandemic has resulted in lower reported number of cases and deaths within the paediatric population, indirect impacts on the health of children in Sub-Saharan Africa such as malnutrition are evident. Data on the socioeconomic factors affecting malnutrition in the under-age population of Sub-Saharan Africa brought by the COVID-19 pandemic remain limited. This paper assesses socioeconomic factors of malnutrition in relation with COVID-19 and potential mitigating measures.

**Methods:** A scoping review of PubMed, Embase, and Web of Science from March 11, 2020, to May 1, 2021, was conducted. The included studies focused on COVID-19, children malnutrition, and Sub-Saharan Africa and adhered to the PRISMA guideline.

**Results:** Among 73 total screened articles, 15 studies filled the inclusion criteria. The identified socioeconomic factors leading to malnutrition in children were reduction in average income or increase in unemployment rate, access to healthcare and food supplements, disrupted food supply chains and increased prices of food products, pauses in humanitarian responses, and reduced access to school-based meals. Potential mitigation measures were food subsidies, food price control measures, the identification of new vulnerable groups and the implementation of financial interventions.

**Conclusion:** Malnutrition amongst Sub-Saharan African children due to COVID-19 is a result of a combination of multiple socioeconomic factors. To stabilize household purchasing power and eventually malnutrition in children in SSA, a combined strategy of initial detection of newly developing vulnerable groups and efficient, rapid financial assistance through mobile phone transfers was suggested. These strategies were proposed in combination with other economical models.

## Background

The indirect, socioeconomic effects of the COVID-19 pandemic have severely impacted the health and malnutrition state of the under-18 population around the globe (Roberton et al., 2020; Parri et al., 2020; Buonsenso et al., 2020, Simba et al., 2020). These effects are anticipated to lead to worse outcomes than the infection itself, making appropriate mitigation strategies essential towards the fight against the pandemic (Roberton et al., 2020; Parri et al., 2020; Buonsenso, 2020, Simba, 2020). Globally, when compared to the adult population, the COVID-19 pandemic resulted in a significantly lower number of cases and an even lower case-fatality rate in the under 18 population (Naja et al., 2020; Coker et al., 2020). Moreover, COVID-19 rarely seemed to end directly in a serious course of disease among children (Ludvigsson, 2020; Parri et al., 2020). Nevertheless, there is a recognizable difference in the direct effect of the pandemic, in terms of lives damaged or lost due to disease and the indirect effect of the pandemic in terms of lives damaged or lost due to economic deprivation (Egger et al., 2021). In this context, effects such as the impairment of current programs to address public health crises such as malnutrition, malaria, HIV, and others, as well as economic effects such as the impairment of family income and the resulting restriction in access to medicine can be highlighted (Coker et al., 2020; Simba et al., 2020). Therefore, especially in low- and middle-income countries (LMICs), among other impacts, the economic downturn resulting from the COVID-19 pandemic is expected to have significant effects on the health and malnutrition of the child population through disturbances in transportation and processing, as well as affordability, limiting access to nutrient-rich foods (El Sadr & Justman, 2020; Fore, et al., 2020; Headey et al., 2020). Unless urgent action is specifically instructed, the pandemic’s indirect effects on child malnutrition may result in a cycle of intergenerational consequences for child growth and cognitive development, with long-term effects on education and chronic disease risk (Da Silva Ferreira et al., 2020). The malnutrition situation in sub-Saharan African (SSA) countries was already in an alarming state before the pandemic, with a heavy burden on the SSA region. According to official data from the World Food Programme (2019), the average percentage of undernourishment in the total population in sub-Saharan countries such as Chad, Zambia, Central African Republic, Rep. of Congo, Uganda, Zimbabwe, as well as Malawi over the years 2016 to 2018 averaged a concerning rate of over 35% (WFP, 2019). Due to COVID-19 the global incidence of malnutrition, specifically the incidence of child wasting, was projected to increase by an alarming 143 %, of which approximately 80% would be projected within SSA and South Asia (Headey et al, 2020). It can be argued that an adaptation of a multidisciplinary perspective through integrating an epidemiological, economic, sociological, and legal viewpoint offers a comprehensive view of the pandemic’s indirect impact. Therefore, the aim of this scientific paper is to discuss a holistic view of the indirect effects of COVID-19 on malnutrition rates in the under-18 population in SSA. Consequently, this scoping review addresses the following two research questions:

1. Through which socioeconomic pathways and to what extent are the effects of the COVID-19 pandemic a triggering factor for an increase in malnutrition in the under-18 population in SSA?
2. What are key measures for mitigating the potential COVID-19 induced rise in malnutrition in the under-18 population in SSA?

## Methods

This scoping review is aligned with the Preferred Reporting Items for Systematic Reviews and Meta-Analyses (PRISMA) Guidelines (Moher et al., 2015). To perform the study at hand, a COVID-19 literature web-scraping tool was applied to allow combined search through common databases such as Pubmed, Embase and Web of Science using predefined syntaxes for identifying relevant literature. Due to the specific focus on the effect of COVID-19 on malnutrition rates in SSA countries, literature addressing predominantly effects on other health determinants or focusing on countries outside SSA were purposely screened out. Additionally, the concerned population was confined to the under-18 age segment. The search syntaxes are composed of terms associated with the COVID-19 pandemic combined with malnutrition by the adaptation of Boolean operators - [OR], [AND], such as “COVID-19” [AND] “Malnutrition” [AND] “sub-Saharan Africa”. Furthermore, the literature review focused on the timeframe from the declaration of the pandemic on 11 March 2020 up to and including 1 May 2021, on which the identification phase of this paper was conducted. This literature review is furthermore focuses only on scientific papers written in English. Following the identification stage of the PRISMA checklist, the results obtained were checked for specific elements such as title and abstract. Thereby the literature is screened and filtered based on the degree to which the publication fits into the framework of the research question. Furthermore, in a third step, the eligibility of the remaining literature was assessed in detail, evaluating and examining the publication in full-text format and hereby applying the inclusion criteria of whether the study is discussing specifically indirect effects of COVID-19 affecting malnutrition among children in SSA. Moreover, the exclusion criteria included the criteria of not addressing countries within SSA, not addressing malnutrition as a co-morbidity rather than malnutrition as a consequence of the COVID-19 pandemic and addressing malnutrition in a different context rather than in relation to COVID-19. For each of the steps within the PRISMA checklist, a data extraction document was created in which the title, the authors, the aim of the study and the central results or findings of the paper, as well as the date of publication and the journal are listed. Subsequently, in the results section, associations were drawn from the data extraction by identifying consistencies as well as distribution of the topics covered between the extracted literature, thereby addressing the research questions.

## Results

The identification phase of the literature selection according to the PRISMA framework resulted in a total of 73 scientific papers. Thereby, 59 papers were identified by database search and a further amount of 14 papers were selected by hand search and snowballing methods. In a second phase, 47 papers were excluded from the identified literature after title and abstract were assessed. After performing a full-article assessment, 15 papers were selected (see Fig. 1; see Annex for exclusion rationale).

**FIGURE 1:**
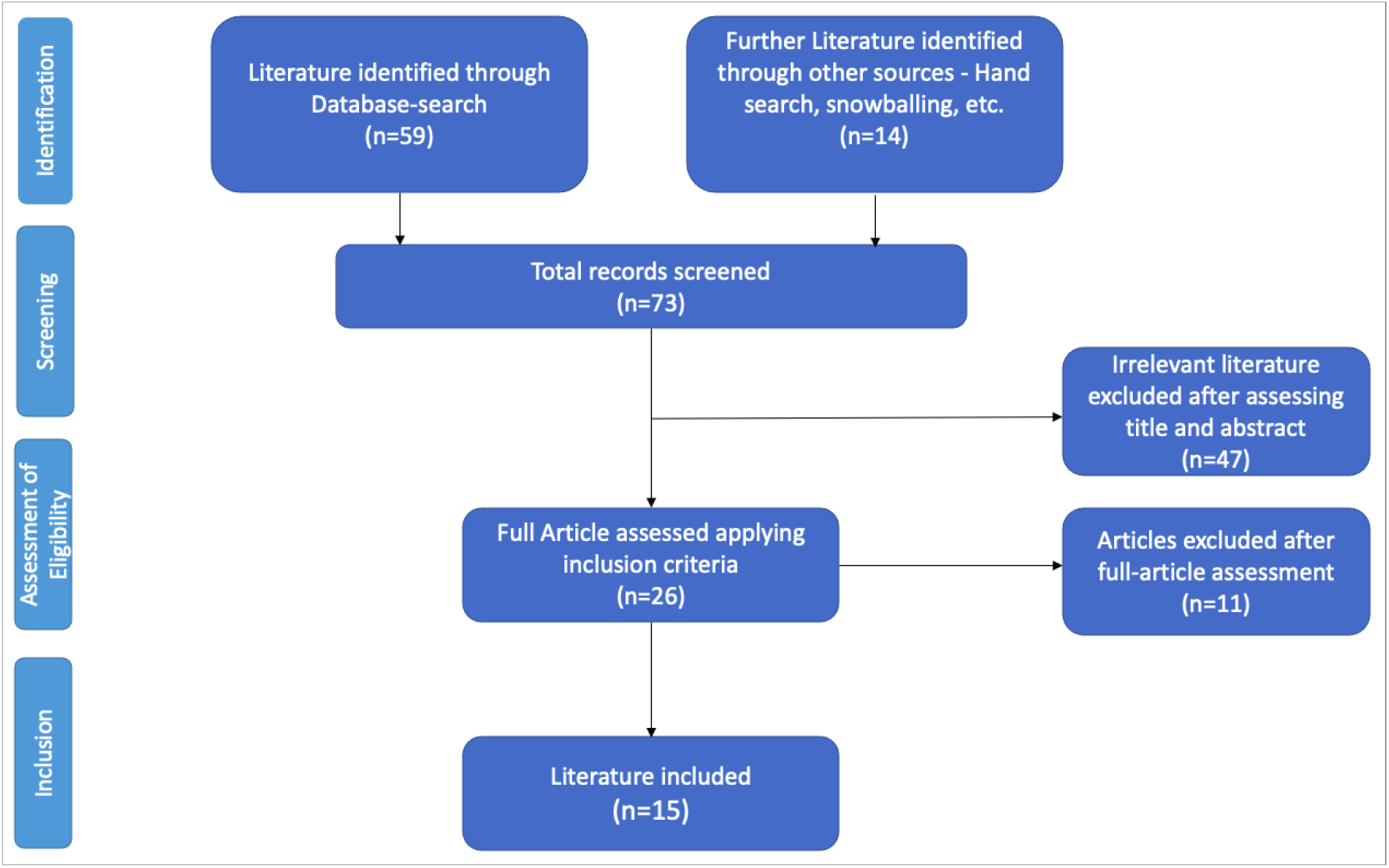
Literature selection in accordance with PRISMA Framework

The most extensive proportion of the selected literature was published between July and September 2020. A decline in the volume of publications in the context of indirect socioeconomic effects of COVID-19 on child-malnutrition within SSA can be observed with the onset of the pandemic up to and including April 2021 (see Fig. 2).

**FIGURE 2:**
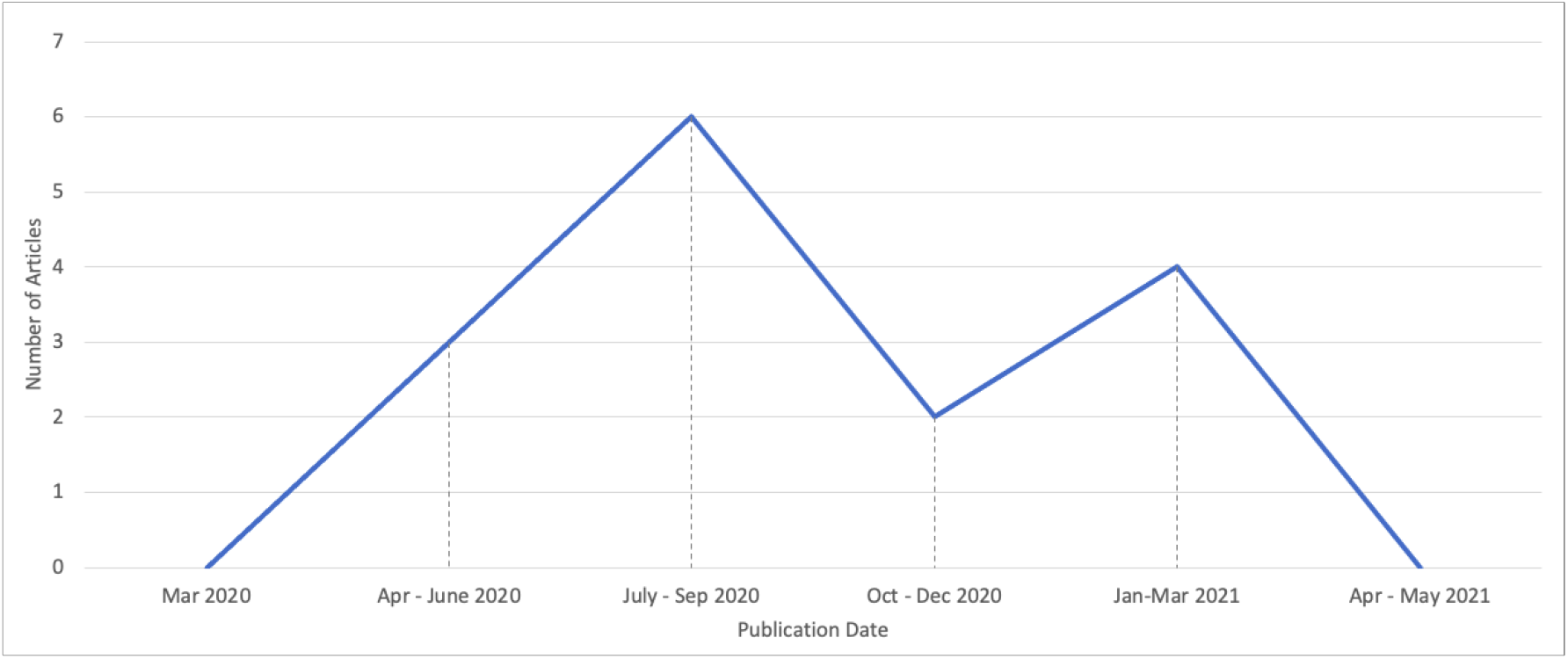
Publication date distribution of the selected literature (n = 15); Source: Own Illustration

The literature consisted of diverse study designs, with almost 70% of the selected literature including a systematic or narrative literature analysis as a core research method, with one of the latter including a meta-analysis. Furthermore, almost 15% of the studies included quantitative online surveys and social accounting matrixes (see Annex). In the following table, the identified socioeconomic factors were considered across the selected literature and listed in detail (see Table 1).

**TABLE 1:** Identified Socioeconomic Factors by Literature Selected

| Main Area of Research and Identified Socioeconomic Factors | Description | Number of Articles |
| --- | --- | --- |
| **Reduction in average income / increasing unemployment** | Control measures leading to an increase in unemployment within the informal economic sector of SSA countries, resulting in reduced household income. | 12 |
| **Access to healthcare and food supplements** | Decrease in access to standard healthcare and food supplements for children who are in pre-stage or already in a state of malnutrition, resulting in an increase in malnutrition rates. | 7 |
| **Disrupted food supply chains and increased prices of staple food products** | Domestic demand being reduced by increased transaction costs and import declines, as well as a reduction in production and sales of nutritious and inexpensive foods resulting in higher prices of staple food products, forcing many families to rely on nutrient-poor alternatives. | 8 |
| **Pauses in humanitarian response through redistribution of support and development funds** | Closure of non-governmental organizations (NGOs), resulting in a decline in often life-saving nutrition services. | 5 |
| **School closures** | Lockdown resulting in the inability for children to have access to school meals, which are a crucial source of adequate nutrition for large parts of under-served children in large parts of SSA. | 6 |

### Reduction in average income / Unemployment

A large part of the selected literature (80%) identified a reduction in average income or a rise in unemployment as one of the factors responsible for increased malnutrition rates in children under 18 years of age as an indirect result of COVID-19 (see Fig. 3). The COVID-19 pandemic caused an undisguised shock to the economies of SSA (Roberton et al., 2020; Headey et al., 2020). In particular, the informal sector of certain SSA-economies, which generates income for many of the region’s families, suffered from the pandemic, making it impossible for large parts of the population to earn a secure income (Govender et al., 2020). According to Govender et al. (2020), the losses in income and the increased unemployment rates resulted from the necessary stringent public health control measures that were disruptive for numerous workers in the informal sector. It can be argued that many businesses in the informal sector released their employees without any period of contract-cancellation, instantly collapsing the hourly wage base and driving the employees and their families and children below the minimum subsistence level. Particularly in countries such as Zimbabwe, which already recorded elevated malnutrition rates in children before the pandemic, the informal employment structure, alongside layoffs in the formal sector, contributed substantially to the rapid worsening of malnutrition rates (Govender et al., 2020).

**FIGURE 3:**
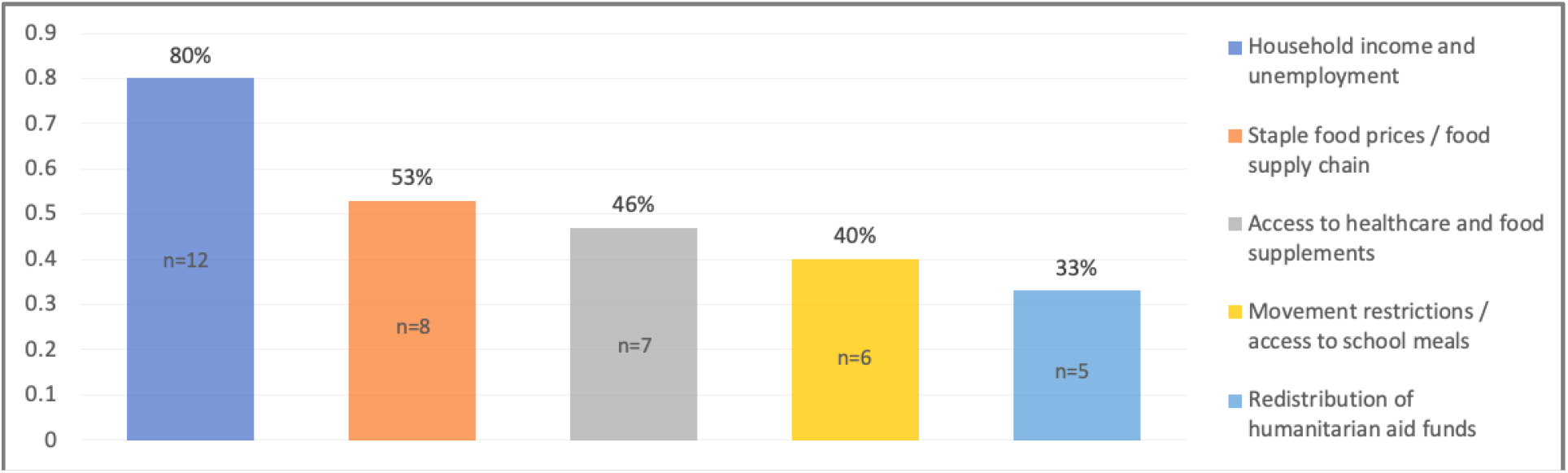
Share of selected literature addressing the identified socio-economic factors; Source: Own Illustration

Households of low level educated workers were particularly affected since such households are especially dependent on a monthly income (Arndt et al., 2020). In the form of a quantitative online survey, Kansiime et al. (2021) conducted a study on COVID-19-induced income shocks among 442 households in Kenya and Uganda. More than two-thirds of the households surveyed reported that diet quality as well as the food security had declined. The results also showed that households dependent on labour income were particularly affected by the effects produced from the pandemic (Kansiime et al., 2021). Consequently, households already living in conditions of uncertainty were especially vulnerable to reduced real incomes (Arndt et al., 2020). Therefore, it can be derived, that children of low level educated households were subjected to increased susceptibility to food insecurity. Egger et al. (2021) conducted a large-scale meta-analysis on the impact of the COVID-19 pandemic on the living standards of, among others, five countries in SSA (Burkina Faso, Ghana, Kenya, Rwanda, and Sierra Leone). The authors observed, among other findings, that a worrying 48% of rural Kenyan households as well as 87% of rural Sierra Leonean households were forced to miss meals or forced to reduce their portion sizes. According to Egger et al. (2021), this exceeds the food insecurity normally present in both observed SSA countries.

Overall, the lower incomes coupled with the increased unemployment rate and the resulting decreased solvency of the local population translated into less accessibility to adequate nutrition, which ultimately led to increased malnutrition and mortality among the under-18 population in SSA (Roberton et al., 2020; Otekunrin et al., 2020). An initial estimate indicated an amount of 47 million children under five years of age affected by malnutrition worldwide prior to the COVID 19 pandemic (Aborode et al., 2020). An additional 6.7 million children are expected to be affected by malnutrition after the first 12 months of the pandemic, 80% of which are living in SSA and South Asia (Aborode et al., 2020).

### Staple food prices / food supply chain

Out of the selected literature, 53% recognized disturbances in the food supply chain and increase in staple food prices as a crucial factor responsible for the elevated malnutrition rates in children under 18 years of age as an indirect result of COVID-19 (see Fig. 3). In this context, disruptions in the food import-export system with negative impacts on local food markets, as well as small-scale enterprises and resulting higher food prices, were identified as key variables (Akseer et al., 2020; Nechifor et al., 2021; Kansiime et al., 2021; Jafri et al., 2021; Aborode et al., 2020). While this effect is less prevalent in well-served urban areas, supply problems and the resulting reduced food availability occurs mainly in rural areas of SSA countries (Akseer et al., 2020). One finding is the declining export rates in SSA countries such as Kenya, negatively impacting the balance of trade and the domestic exchange rate (Nechifor et al., 2021). According to Nechifor et al. (2021), this in turn leads to reduced affordability of imported food and ultimately lower consumption of staple foods, increasing malnutrition of vulnerable groups. Jafri et al. (2021) attributed the increased prices for food mainly to the import side. Due to import declines, the local population was exposed to lower quantities of basic consumer items, resulting in higher prices (Jafri et al., 2021). This was amplified by the lower regional production of food due to limited agricultural activity, translating into even higher prices (Headey et al., 2020; Aborode et al., 2020). As a consequence, this affected the ability of numerous households across Africa to provide adequate food for their children, as they often resorted to cheaper, nutrient-poor alternatives (Aborode et al., 2020; Fore et al., 2020).

### Access to healthcare and food supplements

Approximately half of the selected literature (46%) identified the access to healthcare and food supplements as one of the socioeconomic factors leading to elevated malnutrition rates in children under 18 years of age as a result of COVID-19 (see Fig. 3). Due to conditions such as overcrowded health systems and shifting priorities at the primary care level, access to routine medical care for children with malnutrition or in the pre-state of malnutrition severely worsened (Otenkunrin et al., 2020; Akseer et al., 2020; Ntambara & Chu, 2021; Roberton et al., 2020). According to Akseer et al. (2020), this led to a considerably increase in the risk of child malnutrition, especially if existing conditions persisted over a longer period of time. Due to the treatment of COVID-19 patients, which in some cases completely exhausted the primary healthcare facilities in many SSA countries, basic and regular necessary health services such as the treatment and prevention of child malnutrition through food supplementation were frequently neglected, resulting in increased rates of child malnutrition (Ntambara & Chu, 2021; Aborode et al., 2020, Zar et al., 2020). According to Zar et al. (2020), the severely restricted nutritional programmes in many LMICs indirectly led to increased malnutrition and mortality in the under-18 population. Headey et al. (2020) estimated that COVID-19 led to an additional 128’605 deaths globally in children younger than 5 years through the effect of diminished access to health-care coverage and food supplementation, with 52% of these deaths occurring in SSA.

### Movement restrictions and access to school meals

40% of the selected literature identified movement restrictions and access to school meals as one of the factors responsible for the elevated malnutrition rates in children under 18 years of age as an impact of COVID-19 (see Fig. 3). For many children from low-income households among SSA countries such as Kenya and Uganda, schools were not solely a learning environment, but an environment supplying sufficient nutrition (Kansiime et al., 2021). School meals are one of the few reliable sources of food for many children from low-income households in SSA (Coker et al., 2020, Zar et al., 2020). According to the World Food Programme, 368 million children in pre-primary to secondary school worldwide missed out school meals in 2020, with 148 million of them living in SSA (Coker et al., 2020; World Food Programme (WFP), 2020; WFP & UNICEF, 2020). As a result, a large number of children from households relying on the government’s home-grown school nutrition programme have been exposed to increased malnutrition through the combination of lockdown and school closures (Otekunrin et al., 2020; Coker et al., 2020; Fore et al., 2020; Aborode et al., 2020; Ntambara & Chu, 2021).

### Redistribution of humanitarian aid funds

A relatively narrow share of the literature (33%) also identified the redistribution of humanitarian aid funds as a key component of the increase in malnutrition among the under-18 population in SSA. The reduced activity of numerous non-profit organisations and community centres was further limiting the quality of life and above all, the provision of adequate nutrition for children from low-income households in large parts of SSA (Govender et al., 2020; Fore et al., 2020; Headey et al., 2020; Akseer et al., 2020). This disruption of humanitarian aid, social protection and nutrition services can be attributed to the fact that many states in SSA are forced to reallocate relatively scarce health and social resources to the essential treatment of COVID-19 patients (Fore et al., 2020; Headey et al., 2020).

### Priority areas for action on malnutrition mitigation in the context of COVID-19

After identifying the relevant socio-economic factors affected by COVID-19 and their impact on child malnutrition, the question remains, as to which strategies are potentially promising for counteracting these developments. It is important to emphasize that the identified factors are highly interlinked and therefore subject to potentially having a mutual effect on each other. Due to this possibility, this section thus takes a holistic perspective and examines solutions identified in the selected literature, that are reflected in, or have an impact on several of these factors. Within the literature, strategies regarding food subsidy, with a share of 47% of the literature, food price control measures (26%) and the implementation of financial interventions / financial transfers (53%) were identified. To not go beyond the scope of this paper, the most central strategic measures will be discussed below and a combination of these will be converted into a new combination of measures on the basis of current findings, with the aim of developing an effective response strategy.

The most frequently proposed strategy throughout the selected literature to reduce the COVID-19 induced increase in under-18 malnutrition in SSA, with a literature share of 53% percent of the literature, involved addressing affordability of nutrition through financial interventions. In order to structure these financial transfers and tax exemption measures in the most impactful way possible, it can be argued that, as a first step, it is advisable to combine financial support strategies with the approach proposed by Jafri et al. (2021), of identifying new vulnerable households first. It can be argued that in this way payments could be targeted and those in need can be prioritised for support. This includes workers in the informal sector, small company owners, and private sector employees who have suffered an employment and income loss. Whereas the latter are arguably easier to identify through official numbers, the former proposition would require official public reporting points to identify these groups promptly. Once the new vulnerable groups have been identified, the transfers may subsequently be made using mobile phone transfer donations to ensure payments are made quickly and efficiently (Egger, 2021). Moreover, the approach of Kansiime et al. (2021), which is already applied in large parts of Kenya, may be additionally considered. According to this approach, tax relief is provided instead of a cash transfer if the income falls below subsistence level. In this context, a reduction in the corporation tax is a further possibility, as is a reduction in the value-added tax on small and medium-sized enterprises (Kansiime et al., 2021). It can be argued that reducing personal taxes is expected to increase the household-ability to pay. Reducing small- and medium enterprises (SME) sales taxes can be expected to stabilize corporate losses and increase the job security of employees, ultimately increasing the affordability of food for employee-households and ultimately lead to a decrease in the rate of malnutrition among children under 18 in SSA. According to Nechifor et al. (2021), such fiscal and public spending policies have already contributed to increasing food security during COVID-19, particularly in urban areas. However, it should be noted that the beneficiary pools should be sufficiently extended to rural areas, as well as to the poorest income percentiles, to allow for a uniform effect across all beneficiaries (Nechifor et al., 2021). Moreover, as one major cause of the rise in malnutrition among under-18s in SSA identified in this paper is the increase in unemployment, an additional measure would be the improvement in access to legal protection and courts, addressing the identified need of higher employee protection in the informal sector (Coker et al., 2020). One potential rationale lies in governments ensuring that employers are obliged to provide the employee with a formal employment contract with a corresponding notice period and/or reduced working hours including salary. This transfer to a more formal working relationship would specifically counteract the fact that households of informal workers are rapidly exposed to malnutrition in the event of employment loss, thus creating a legally authenticated and hence better protected employment relationship.

## Discussion

Regarding the first research question, the scoping review indicated that an increase in the malnutrition rates of children in SSA due to the COVID-19 pandemic is evident and clearly measurable based on current estimates. The selected literature identified the effect of increased unemployment and lower income as a key indicator, as 80% of the literature reviewed found this factor to be crucial. Although this study has subdivided the individual sub-areas based on indepth analysis of the selected literature, it is important to recognize that the impact of COVID-19 on child malnutrition rates in SSA occurs in an indirect and interconnected manner and involves interaction of a set of factors.

Other factors identified across the selected literature consisted of higher staple food prices, interruptions in the food supply chain, reduced access to school-based meals, diminished access to healthcare and food assistance, as well as the interrupted activity of humanitarian aid in malnutrition mitigation. It can be argued that these most likely interacted strongly with each other, as well as with the effect of increased unemployment and reduced incomes in SSA. It can be argued that consequently, throughout the literature reviewed, there was a perceived need for a multidisciplinary approach to limit the negative impact of COVID-19 on child malnutrition in SSA. Therefore, it can additionally be interpreted that intersectoral cooperation between different stakeholders such as government ministries like health promotion and economic- and development promotion, international NGOs and hospitals is central to counteracting this effect. One limitation of this paper is that while most studies focused entirely on the SSA region, some studies, while focusing on SSA, observed additional LMICs besides LMICs in SSA. This paper aimed to specifically extract the findings from the SSA region. Nevertheless, this factor could have potentially led to minor interpretation biases.

## Conclusion

To sum up, this scoping review aimed to identify the most relevant socio-economic pathways of COVID-19, leading to a potential increase in malnutrition in the unter-18 population in SSA and to elaborate urgently needed mitigation strategies. The overall findings indicate an increase in the rate of malnutrition among the under-18 population due to the COVID-19 pandemic in the SSA region, mainly as a result of increased unemployment and reduced incomes. Furthermore, effects such as disturbances in the food supply chain and increased staple food prices, access to healthcare and nutritional supplements, access to school-based meals, as well as the paused humanitarian aid activities were factors that lead to this development. Moreover, the objective of this study was to synthesize possible strategies from existing literature and combine them where possible to develop novel strategies to address the increased malnutrition rate. A combined approach of initial identification of newly emerging vulnerable groups and efficient, rapid financial support to vulnerable households to stabilise household purchasing power and ultimately malnutrition in children in SSA was proposed.

This assistance is to be supported by a reduction in personal taxes for people living in vulnerable households to further strengthen purchasing power. In addition, a reduction in the value-added tax in SMEs in SSA countries was considered to stabilize the labour market and maintain job security as a means to secure household income for child malnutrition prevention. As a final measure, this paper discussed the solution that governments should be responsible for establishing more formal employment conditions to enable a strengthened employment relationship that provides more rights to the worker and arguably less poverty traps and child malnutrition in households.

In future studies, it is crucial to address not only the key factors identified here, but also their interactions and mutual impacts in order to draw further conclusions on appropriate mitigation strategies for COVID-19-induced increases in malnutrition in SSA countries.

## Supporting information

Supplementary File 1 - Search Strategy Syntax

Supplementary File 2 - Literature List Database Search

Supplemental File 3 - Literature Snowball and Hand Search

Supplementary File 4 - Literature Screening and Assessment of Eligibility

Supplementary File 5 - Selected Literature - Addressed Areas of Research and Study Designs

## Data Availability

All studies and data supporting the results can be found in the publicly available datasets of PubMed, Embase, and Web of Science.

## Author Contribution

PS and SRV conceived and designed the study. OK, and EO made substantial contributions in reviewing the design of the study. PS and EO helped with screening of literature in databases. PS conducted the literature review. PS and SRV drafted the manuscript. OK and EO contributed by revising the manuscript critically for important intellectual content. PS, SRV, OK, and EO critically reviewed the manuscript. All authors contributed to final approval of the version to be submitted.

## Ethical Approval

Data were collected from publicly available datasets, and no ethical approval was needed.

