## Supplementary File 1 - Search Strategy Syntax for "The COVID-19 pandemic and child malnutrition in sub-Saharan Africa: A scoping review"

### Additional File 1 – Search Strategy

| Databases | Search Syntax [Title/Abstract] |
|---|---|
| Covid-19 Literature-Clustering Tool - PubMed, Medline, etc. | ('coronavirus' OR 'covid-19' OR 'covid' OR 'sars2' OR 'sars-cov' OR 'sars-cov-2' OR 'ncov' OR 'cov2' OR 'severe acute respiratory pneumonia outbreak' OR 'coronaviridae' OR 'corona virus‘)<br>AND<br>('children' OR 'child' OR 'infant' OR 'infants' OR 'pediatric' OR 'paediatric' OR 'nursery' OR 'kindergarten' OR 'kid' OR 'kids' OR 'toddler' OR 'toddlers' OR 'adolescent' OR 'adolescents' OR 'babies' OR ‚baby')<br>AND<br>('Africa' OR 'africa' OR 'SSA' OR 'ssa' OR 'SSA countries' OR 'Angola' OR 'Burundi' OR 'Congo' OR 'Cameroon' OR 'Chad' OR 'Guinea' OR 'Gabon' OR 'Kenya' OR 'Nigeria' OR 'Rwanda' OR 'Sao' OR 'Tanzania' OR 'Uganda' OR 'Sudan' OR 'Djibouti' OR 'Eritrea' OR 'Ethiopia' OR 'Somalia' OR 'Botswana' OR 'Comoros' OR 'Lesotho' OR 'Madagascar' OR 'Malawi' OR 'Mauritius' OR 'Mozambique' OR 'Namibia' OR 'Seychelles' OR 'Swaziland' OR 'Zambia' OR 'Zimbabwe' OR 'Benin' OR 'Mali' OR 'Burkina' OR 'Verde' OR 'Ivory' OR 'Gambia' OR 'Ghana' OR 'Liberia' OR 'Mauritania' OR 'Niger' OR 'Senegal' OR 'Leone' OR ‚Togo')<br>AND<br>('malnutrition' OR 'undernourishment' OR 'hunger' OR 'famine' OR 'starvation' OR 'poor diet' OR 'lack of food' OR 'malnourishment' OR 'inadequate diet' OR 'stunting' OR 'wasting' OR 'undernutrition' OR ‘food systems’ OR ‘household-income’ OR ‘food security’ OR ‘food insecurity’ OR ‘food affordability’ OR ‘food accessibility’) |
