## Supplementary File 2 - Literature List Database Search for "The COVID-19 pandemic and child malnutrition in sub-Saharan Africa: A scoping review"

### Database Search (n=59)

| Title | Authors | Date | Journal | URL | Abstract |
|---|---|---|---|---|---|
| COVID-19 and Pediatric Lung Disease: A South African Tertiary Center Experience | Gray, Diane M.; Davies, Mary-Ann; Githinji, Leah; Levin, Michael; Mapani, Muntanga; Nowalaza, Zandiswa; Washaya, Norbertta; Yassin, Aamir; Zampoli, Marco; Zar, Heather J.; Vanker, Aneesa | 2021-01-20 | Front Pediatr | https://www.ncbi.nlm.nih.gov/pubmed/33553073/; https://doi.org/10.3389/fped.2020.614076 | The COVID-19 pandemic led to rapid global spread with far-reaching impacts on health-care systems. Whilst pediatric data consistently shown a milder disease course, chronic lung disease has been identified as a risk factor for hospitalization and severe disease. In Africa, comprised predominantly of low middle-income countries (LMIC), the additional burden of HIV, tuberculosis, malnutrition and overcrowding is high and further impacts health risk. This paper reviewed the literature on COVID-19 and chronic lung disease in children and provides our experience from an African pediatric pulmonary center in Cape Town, South Africa. South African epidemiological data confirms a low burden of severe disease with children <18 years comprising 8% of all diagnosed cases and 3% of all COVID-19 admissions. A decrease in hospital admission for other viral lower respiratory tract infections was found. While the pulmonology service manages children with a wide range of chronic respiratory conditions including bronchiectasis, cystic fibrosis, asthma, interstitial lung disease and children with tracheostomies, no significant increase in COVID-19 admissions were noted and in those who developed COVID-19, the disease course was not severe. Current evidence suggests that pre-existing respiratory disease in children does not appear to be a significant risk factor for severe COVID-19. Longitudinal data are still needed to assess risk in children with immunosuppression and interstitial lung diseases. The indirect impacts of the pandemic response on child respiratory health are notable and still likely to be fully realized and quantified. Ensuring children have access to full preventive and care services during this time is priority. |
| Kidney diseases and COVID-19 infection: | Askari, Hassan; Sanadgol, Nima; Azarnezhad, Asaad; Tajbakhsh, Amir; Rafiei, | 2021-01-20 | Heliyon | https://doi.org/10.1016/j.heliyon.2021.e06008; https://api.elsevier.co | Recently, the novel coronavirus disease 2019 (COVID-19), has attracted the attention of scientists where it has a high mortality rate among older adults and individuals suffering from chronic diseases, such as chronic kidney diseases (CKD). It is important to elucidate molecular mechanisms by which COVID-19 |

| Title | Authors | Date | Journal | URL | Abstract |
|---|---|---|---|---|---|
| causes and effect, supportive therapeutics and nutritional perspectives | Hossein; Safarpour, Ali Reza; Gheibihayat, Seyed Mohammad; Raeis-Abdollahi, Ehsan; Savardashtaki, Amir; Ghanbariasad, Ali; Omidifar, Navid | | | m/content/article/pii/S2405844021001134; https://www.ncbi.nlm.nih.gov/pubmed/33495739/; https://www.sciencedirect.com/science/article/pii/S2405844021001134 | affects the kidneys and accordingly develop proper nutritional and pharmacological strategies. Although numerous studies have recently recommended several approaches for the management of COVID-19 in CKD, its impact on patients with renal diseases remains the biggest challenge worldwide. In this paper, we review the most recent evidence regarding causality, potential nutritional supplements, therapeutic options, and management of COVID-19 infection in vulnerable individuals and patients with CKD. To date, there is no effective treatment for COVID-19-induced kidney dysfunction, and current treatments are yet limited to anti-inflammatory (e.g. ibuprofen) and anti-viral medications (e.g. Remdesivir, and Chloroquine/Hydroxychloroquine) that may increase the chance of treatment. In conclusion, the knowledge about kidney damage in COVID-19 is very limited, and this review improves our ability to introduce novel approaches for future clinical trials for this contiguous disease. |
| A highly pathogenic GI-19 lineage infectious bronchitis virus originated from multiple recombination events with broad tissue tropism | Hou, Yutong; Zhang, Lili; Ren, Mengting; Han, Zongxi; Sun, Junfeng; Zhao, Yan; Liu, Shengwang | 2020-05-04 | Virus Res | https://www.sciencedirect.com/science/article/pii/S016817022030191X?v=s5; https://doi.org/10.1016/j.virusres.2020.198002; https://www.ncbi.nlm.nih.gov/pubmed/32380209/; https://api.elsevier.com/content/article/pii/S016817022030191X | Abstract In the present study, an IBV strain I0305/19 was isolated from a diseased commercial broiler flock in 2019 in China with high morbidity and mortality. The isolate I0305/19 was clustered together with viruses in sublineage D of GI-19 lineage on the basis of the complete S1 sequence analysis. Isolate I0305/19 and other GI-19 viruses isolated in China have the amino acid sequence MIA at positions 110 -112 in the S protein. Further analysis based on the complete genomic sequence showed that the isolate emerged through at least four recombination events between GI-19 ck/CH/LJS/120848- and GI-13 4/91-like strains, in which the S gene was found to be similar to that of the GI-19 ck/CH/LJS/120848-like strain. Pathological assessment showed the isolate was a nephropathogenic IBV strain that caused high morbidity of 100% and mortality of 80% in 1-day-old specific-pathogen-free (SPF) chicks. The isolate I0305/19 exhibited broader tropisms in different tissues, including tracheas, lungs, bursa of Fabricius, spleen, liver, kidneys, proventriculus, small intestines, large intestines, cecum, and cecal tonsils. Furthermore, subpopulations of the challenge virus were found in tissues of infected chickens; this finding is important in understanding how the virulent IBV strains can potentially replicate |

| Title | Authors | Date | Journal | URL | Abstract |
|---|---|---|---|---|---|
| | | | | | and evolve to cause disease. This information is also valuable for understanding the mechanisms of replication and evolution of other coronaviruses such as the newly emerged SARS-CoV-2. |
| COVID-19: a novel menace for the practice of nephrology and how to manage it with minor devastation? | Ulu, Sena; Gungor, Ozkan; Gok Oguz, Ebru; Hasbal, Nuri Baris; Turgut, Didem; Arici, Mustafa | 2020-07-27 | Renal failure | https://doi.org/10.1080/0886022x.2020.1797791; https://www.ncbi.nlm.nih.gov/pubmed/32713282/ | Coronavirus disease 19 (COVID-19) became a nightmare for the world since December 2019. Although the disease affects people at any age; elderly patients and those with comorbidities were more affected. Everyday nephrologists see patients with hypertension, chronic kidney disease, maintenance dialysis treatment or kidney transplant who are also high-risk groups for the COVID-19. Beyond that, COVID-19 or severe acute respiratory syndrome (SARS) due to infection may directly affect kidney functions. This broad spectrum of COVID-19 influence on kidney patients and kidney functions obviously necessitate an up to date management policy for nephrological care. This review overviews and purifies recently published literature in a question to answer format for the practicing nephrologists that will often encounter COVID-19 and kidney related cases during the pandemic times. |
| COVID-19 in children: what did we learn from the first wave? | Bogiatzopoulou, Aliki; Mayberry, Huw; Hawcutt, Daniel B.; Whittaker, Elizabeth; Munro, Alasdair; Roland, Damian; Simba, Justus; Gale, Christopher; Felsenstein, Susanna; Abrams, Elissa; Jones, Caroline B.; Lewins, Ian; Rodriguez-Martinez, Carlos R.; Fernandes, Ricardo M.; Stilwell, Philippa A.; Swann, | 2020-09-18 | Paediatr Child Health (Oxford) | https://doi.org/10.1016/j.paed.2020.09.005; https://www.sciencedirect.com/science/article/pii/S1751722220301591?v=s5; https://www.ncbi.nlm.nih.gov/pubmed/32983255/; https://api.elsevier.com/content/article/pii/S1751722220301591 | A pandemic caused by the novel coronavirus, severe acute respiratory syndrome - coronavirus 2 (SARS-CoV-2), has caused high rates of mortality, predominantly in adults. Children are significantly less affected by SARS-CoV-2 with far lower rates of recorded infections in children compared to adults, milder symptoms in the majority of children and very low mortality rates. A suspected late manifestation of the disease, paediatric inflammatory multisystem syndrome - temporally associated with SARS-CoV-2 (PIMS-TS), has been seen in small numbers of children and has a more severe disease course than acute SARS-CoV-2. The pandemic has meant that children around the world have been kept off school, isolated from their extended family and friends and asked to stay inside. The UK has just been declared as being in an economic recession and unemployment rates are increasing. These indirect effects of SARS-CoV-2 are likely to have a significant impact on many children for years to come. Consolidating the knowledge that has accumulated during the first wave of this pandemic is essential for recognising the clinical signs, symptoms and effective |

| Title | Authors | Date | Journal | URL | Abstract |
|---|---|---|---|---|---|
| | Olivia; Bhopal, Sunil; Sinha, Ian; Harwood, Rachel | | | | treatment strategies for children; identifying children who may be at increased risk of severe SARS-CoV-2 infection; planning the safe delivery of healthcare and non-health related services that are important for childrens' wellbeing; and engaging in, and developing, research to address the things that are not yet known. This article summarises the evidence that has emerged from the early phase of the pandemic and offers an overview for those looking after children or planning services. |
| The differential impact of pediatric COVID-19 between high-income countries and low- and middle-income countries: A systematic review of fatality and ICU admission in children worldwide | Kitano, Taito; Kitano, Mao; Krueger, Carsten; Jamal, Hassan; Al Rawahi, Hatem; Lee-Krueger, Rachelle; Sun, Rose Doulin; Isabel, Sandra; García-Ascaso, Marta Taida; Hibino, Hiromi; Camara, Bettina; Isabel, Marc; Cho, Leanna; Groves, Helen E.; Piché-Renaud, Pierre-Philippe; Kossov, Michael; Kou, Ikuho; Jon, Ilsu; Blanchard, Ana C.; Matsuda, Nao; Mahood, Quenby; Wadhwa, Anupma; Bitnun, Ari; Morris, Shaun K. | 2021-01-29 | PLoS One | https://doi.org/10.1371/journal.pone.0246326; https://www.ncbi.nlm.nih.gov/pubmed/33513204/ | BACKGROUND: The overall global impact of COVID-19 in children and regional variability in pediatric outcomes are presently unknown. METHODS: To evaluate the magnitude of global COVID-19 death and intensive care unit (ICU) admission in children aged 0–19 years, a systematic review was conducted for articles and national reports as of December 7, 2020. This systematic review is registered with PROSPERO (registration number: CRD42020179696). RESULTS: We reviewed 16,027 articles as well as 225 national reports from 216 countries. Among the 3,788 global pediatric COVID-19 deaths, 3,394 (91.5%) deaths were reported from low- and middle-income countries (LMIC), while 83.5% of pediatric population from all included countries were from LMIC. The pediatric deaths/1,000,000 children and case fatality rate (CFR) were significantly higher in LMIC than in high-income countries (HIC) (2.77 in LMIC vs 1.32 in HIC; $p < 0.001$ and 0.24% in LMIC vs 0.01% in HIC; $p < 0.001$, respectively). The ICU admission/1,000,000 children was 18.80 and 1.48 in HIC and LMIC, respectively ($p < 0.001$). The highest deaths/1,000,000 children and CFR were in infants < 1 year old (10.03 and 0.58% in the world, 5.39 and 0.07% in HIC and 10.98 and 1.30% in LMIC, respectively). CONCLUSIONS: The study highlights that there may be a larger impact of pediatric COVID-19 fatality in LMICs compared to HICs. |
| Evaluation of patient characteristic | Mash, Robert James; Presence-Vollenhoven, Mellisa; Adeniji, | 2021-01-26 | BMJ Open | https://www.ncbi.nlm.nih.gov/pubmed/33500292/; | OBJECTIVES: To describe the characteristics, clinical management and outcomes of patients with COVID-19 at district hospitals. DESIGN: A descriptive observational cross-sectional study. SETTING: District hospitals (4 |

| Title | Authors | Date | Journal | URL | Abstract |
|---|---|---|---|---|---|
| s, management and outcomes for COVID-19 at district hospitals in the Western Cape, South Africa: descriptive observational study | Adeloye; Christoffels, Renaldo; Doubell, Karlien; Eksteen, Lawson; Hendrikse, Amee; Hutton, Lauren; Jenkins, Louis; Kapp, Paul; Lombard, Annie; Marais, Heleen; Rossouw, Liezel; Stuve, Katrin; Ugoagwu, Abi; Williams, Beverley | | | https://doi.org/10.1136/bmjopen-2020-047016 | in metro and 4 in rural health services) in the Western Cape, South Africa. District hospitals were small (<150 beds) and led by family physicians. PARTICIPANTS: All patients who presented to the hospitals' emergency centre and who tested positive for COVID-19 between March and June 2020. PRIMARY AND SECONDARY OUTCOME MEASURES: Source of referral, presenting symptoms, demographics, comorbidities, clinical assessment and management, laboratory turnaround time, clinical outcomes, factors related to mortality, length of stay and location. RESULTS: 1376 patients (73.9% metro, 26.1% rural). Mean age 46.3 years (SD 16.3), 58.5% females. The majority were self-referred (71%) and had comorbidities (67%): hypertension (41%), type 2 diabetes (25%), HIV (14%) and overweight/obesity (19%). Assessment of COVID-19 was mild (49%), moderate (18%) and severe (24%). Test turnaround time (median 3.0 days (IQR 2.0–5.0 days)) was longer than length of stay (median 2.0 day (IQR 2.0–3.0)). The most common treatment was oxygen (41%) and only 0.8% were intubated and ventilated. Overall mortality was 11%. Most were discharged home (60%) and only 9% transferred to higher levels of care. Increasing age (OR 1.06 (95% CI 1.04 to 1.07)), male (OR 2.02 (95% CI 1.37 to 2.98)), overweight/obesity (OR 1.58 (95% CI 1.02 to 2.46)), type 2 diabetes (OR 1.84 (95% CI 1.24 to 2.73)), HIV (OR 3.41 (95% CI 2.06 to 5.65)), chronic kidney disease (OR 5.16 (95% CI 2.82 to 9.43)) were significantly linked with mortality (p<0.05). Pulmonary diseases (tuberculosis (TB), asthma, chronic obstructive pulmonary disease, post-TB structural lung disease) were not associated with increased mortality. CONCLUSION: District hospitals supported primary care and shielded tertiary hospitals. Patients had high levels of comorbidities and similar clinical pictures to that reported elsewhere. Most patients were treated as people under investigation. Mortality was comparable to similar settings and risk factors identified. |
| A review of properties, nutritional | Meireles, Diana; Gomes, João; Lopes, Lara; | 2020-08-17 | ADV TRADIT | https://www.ncbi.nlm.nih.gov/pmc/articles/PMC7430547/ | Moringa oleifera L. from the Moringaceae family is a perennial tree widely cultivated in many tropic regions and easily grown even in adverse conditions. M. oleifera is also known as the miracle tree, which for centuries has been |

| Title | Authors | Date | Journal | URL | Abstract |
| --- | --- | --- | --- | --- | --- |
| and pharmaceutical applications of Moringa oleifera: integrative approach on conventional and traditional Asian medicine | Hinzmann, Mariana; Machado, Jorge | | MED (ADTM) | | indicated for traditional medicine. With no reports of side effects, in doses achievable by ingestion, different parts of M. oleifera is used to treat several conditions, such as malnutrition, diabetes, blindness, anemia, hypertension, stress, depression, skin, arthritis, joints and kidney stones disorders. This plant also showed capacity of helping in maintenance of the cardiovascular system health, blood-glucose levels and providing anti-oxidant, anti-inflammatory and anti-cancer activity as well as the regulation of urinary tract and lactation in nursing women. The seed and leaves powder has water purification properties through flocculation. It also supplements the food in the human diet and in the fortification of livestock feed, especially in developing countries. So, M. oleifera properties have also been applied to cosmetic and byproducts industries due to the high nutritive and protective properties of its seed oil. According to the holistic or traditional medicine, M. oleifera has very relevant therapeutic properties and applications depending on the constitution, somatic and psychological needs of patients. It is usually referred as a natural product that can treat different physical and psychological health aspects, offering an energetic action and structural rebuilder of the body and promoting emotions of highly positive attitudes towards life. The high and specific immunological potential of M. oleifera leads us to suggest an in-depth study to assess the hypothesis of conferring a supportive effect against Covid-19 disease. |
| COVID-19: Review of a 21st Century Pandemic from Etiology to Neuro-psychiatric Implications | Yamamoto, Vicky; Bolanos, Joe F.; Fiallos, John; Strand, Susanne E.; Morris, Kevin; Shahrokhinia, Sanam; Cushing, Tim R.; Hopp, Lawrence; Tiwari, Ambooj; Hariri, Robert; Sokolov, Rick; Wheeler, Christopher; Kaushik, | 2020-09-15 | Journal of Alzheimer's disease : JAD | https://doi.org/10.3233/jad-200831; https://www.ncbi.nlm.nih.gov/pubmed/32925078/ | COVID-19 is a severe infectious disease that has claimed >150,000 lives and infected millions in the United States thus far, especially the elderly population. Emerging evidence has shown the virus to cause hemorrhagic and immunologic responses, which impact all organs, including lungs, kidneys, and the brain, as well as extremities. SARS-CoV-2 also affects patients', families', and society's mental health at large. There is growing evidence of re-infection in some patients. The goal of this paper is to provide a comprehensive review of SARS-CoV-2-induced disease, its mechanism of infection, diagnostics, therapeutics, and treatment strategies, while also focusing on less attended aspects by previous studies, including nutritional support, psychological, and rehabilitation of the |

| Title | Authors | Date | Journal | URL | Abstract |
|---|---|---|---|---|---|
| | Ajeet; Elsayegh, Ashraf; Eliashiv, Dawn; Hedrick, Rebecca; Jafari, Behrouz; Johnson, J. Patrick; Khorsandi, Mehran; Gonzalez, Nestor; Balakhani, Guita; Lahiri, Shouri; Ghavidel, Kazem; Amaya, Marco; Kloor, Harry; Hussain, Namath; Huang, Edmund; Cormier, Jason; Wesson Ashford, J.; Wang, Jeffrey C.; Yaghobian, Shadi; Khorrami, Payman; Shamloo, Bahman; Moon, Charles; Shadi, Payam; Kateb, Babak | | | | pandemic and its management. We performed a systematic review of >1,000 articles and included 425 references from online databases, including, PubMed, Google Scholar, and California Baptist University's library. COVID-19 patients go through acute respiratory distress syndrome, cytokine storm, acute hypercoagulable state, and autonomic dysfunction, which must be managed by a multidisciplinary team including nursing, nutrition, and rehabilitation. The elderly population and those who are suffering from Alzheimer's disease and dementia related illnesses seem to be at the higher risk. There are 28 vaccines under development, and new treatment strategies/protocols are being investigated. The future management for COVID-19 should include B-cell and T-cell immunotherapy in combination with emerging prophylaxis. The mental health and illness aspect of COVID-19 are among the most important side effects of this pandemic which requires a national plan for prevention, diagnosis and treatment. |
| Coronaviruses: An Updated Overview of Their Replication and Pathogenesis | Wang, Yuhang; Grunewald, Matthew; Perlman, Stanley | 2020-08-25 | Coronaviruses | https://www.ncbi.nlm.nih.gov/pubmed/32833200/; https://doi.org/10.1007/978-1-0716-0900-2_1 | Coronaviruses (CoVs), enveloped positive-sense RNA viruses, are characterized by club-like spikes that project from their surface, an unusually large RNA genome, and a unique replication strategy. CoVs cause a variety of diseases in mammals and birds ranging from enteritis in cows and pigs, and upper respiratory tract and kidney disease in chickens to lethal human respiratory infections. Most recently, the novel coronavirus, SARS-CoV-2, which was first identified in Wuhan, China in December 2019, is the cause of a catastrophic pandemic, COVID-19, with more than 8 million infections diagnosed worldwide by mid-June 2020. Here we provide a brief introduction to CoVs discussing their replication, pathogenicity, and current prevention and treatment strategies. We |

| Title | Authors | Date | Journal | URL | Abstract |
|---|---|---|---|---|---|
| | | | | | will also discuss the outbreaks of the highly pathogenic Severe Acute Respiratory Syndrome Coronavirus (SARS-CoV) and Middle Eastern Respiratory Syndrome Coronavirus (MERS-CoV), which are relevant for understanding COVID-19. |
| Lessons learned from the A (H1N1) Influenza Pandemic | Vousden, Nicola; Knight, Marian | 2020-10-12 | Best Pract Res Clin Obstet Gynaecol | https://www.sciencedirect.com/science/article/pii/S1521693420301577?v=s5; https://api.elsevier.com/content/article/pii/S1521693420301577; https://www.ncbi.nlm.nih.gov/pubmed/33144076/; https://doi.org/10.1016/j.bpobgyn.2020.08.006 | Influenza in pregnancy is a common condition that is associated with increased risk of hospital admission. Women with comorbidities are at greater risk of severe outcomes. There are substantial gaps in our knowledge of the impact of severe influenza on perinatal outcomes, especially in low and middle-income countries, but preterm birth, fetal death, infant respiratory infection and hospital admission may be increased. Thus, influenza is major burden on health services. Immunisation is cost-effective, safe and effective at preventing influenza in pregnant women and their infants but policies and uptake vary worldwide. Operational challenges and concern over the safety, efficacy and necessity of the immunisation are common and there is a lack of evidence on how to overcome these barriers. This review identifies learning points relevant to the current COVID-19 pandemic through describing the epidemiology and impact of seasonal and A(H1N1)pdm09 influenza in pregnancy, alongside the effectiveness and use of immunisation. |
| Impact of Vaccines; Health, Economic and Social Perspectives | Rodrigues, Charlene M. C.; Plotkin, Stanley A. | 2020-07-14 | Front Microbiol | https://www.ncbi.nlm.nih.gov/pubmed/32760367/; https://doi.org/10.3389/fmicb.2020.01526 | In the 20th century, the development, licensing and implementation of vaccines as part of large, systematic immunization programs started to address health inequities that existed globally. However, at the time of writing, access to vaccines that prevent life-threatening infectious diseases remains unequal to all infants, children and adults in the world. This is a problem that many individuals and agencies are working hard to address globally. As clinicians and biomedical scientists we often focus on the health benefits that vaccines provide, in the prevention of ill-health and death from infectious pathogens. Here we discuss the health, economic and social benefits of vaccines that have been identified and studied in recent years, impacting all regions and all age groups. After learning of the emergence of SARS-CoV-2 virus in December 2019, and its potential for global dissemination to cause COVID-19 disease was realized, there was an urgent need to develop vaccines at an unprecedented rate and scale. As we |

| Title | Authors | Date | Journal | URL | Abstract |
|---|---|---|---|---|---|
| | | | | | appreciate and quantify the health, economic and social benefits of vaccines and immunization programs to individuals and society, we should endeavor to communicate this to the public and policy makers, for the benefit of endemic, epidemic, and pandemic diseases. |
| The Critical Need for Pooled Data on COVID-19 in African Children: An AFREhealth Call for Action through Multi-Country Research Collaboration | Sam-Agudu, Nadia A; Rabie, Helena; Pipo, Michel Tshiasuma; Byamungu, Liliane Nsuli; Masekela, Refiloe; van der Zalm, Marieke M; Redfern, Andrew; Dramowski, Angela; Mukalay, Abdon; Gachuno, Onesmus W; Mongweli, Nancy; Kinuthia, John; Ishoso, Daniel Katuashi; Amoako, Emmanuella; Agyare, Elizabeth; Agbeno, Evans K; Jibril, Aishatu Mohammed; Abdullahi, Asara M; Amadi, Oma; Umar, Umar Mohammed; Ayele, Birhanu T; Machekano, Rhoderick N; Nyasulu, Peter S; Hermans, Michel P; Otshudiema, John Otokoye; Bongo-Pasi | 2021-02-13 | Clin Infect Dis | https://doi.org/10.1093/cid/ciab142; https://www.ncbi.nlm.nih.gov/pubmed/33580256/ | Globally, there are prevailing knowledge gaps in the epidemiology, clinical manifestations, and outcomes of SARS-CoV-2 infection among children and adolescents; however, these gaps are especially wide in African countries. The availability of robust age-disaggregated data is a critical first step in improving knowledge on disease burden and manifestations of COVID-19 among children. Furthermore, it is essential to improve understanding of SARS-CoV-2 interactions with comorbidities and co-infections such as HIV, tuberculosis, malaria, sickle cell disease and malnutrition, which are highly prevalent among children in sub-Saharan Africa. The African Forum for Research and Education in Health (AFREhealth) COVID-19 Research Collaboration on Children and Adolescents is conducting studies across Western, Central, Eastern, and Southern Africa to address existing knowledge gaps. This consortium is expected to generate key evidence to inform clinical practice and public health policymaking for COVID-19, while concurrently addressing other major diseases affecting children in African countries. |

| Title | Authors | Date | Journal | URL | Abstract |
| --- | --- | --- | --- | --- | --- |
| | Nswe, Christian; Kayembe, Jean-Marie N; Mbala-Kingebeni, Placide; Muyembe-Tamfum, Jean-Jacques; Aanyu, Hellen Tukamuhebwa; Musoke, Philippa; Fowler, Mary Glenn; Sewankambo, Nelson; Suleman, Fatima; Adejumo, Prisca; Tsegaye, Aster; Mteta, Alfred; Noormahomed, Emilia V; Deckelbaum, Richard J; Zumla, Alimuddin; Mavungu Landu, Don Jethro; Tshilolo, Léon; Zigabe, Serge; Goga, Ameena; Mills, Edward J; Umar, Lawal W; Kruger, Mariana; Mofenson, Lynne M; Nachega, Jean B | | | | |
| COVID-19 Management In Pediatrics | Alcindor, Magalie L.; Alcindor, FitzGerald; Richard, Kristy E.; Ajay, Geetha; Denis, Anne Marie; Dickson, Darlene M.; Lawal, Ekaete; | 2021-03-09 | J Nurse Pract | https://api.elsevier.com/content/article/pii/S1555415521000660; https://doi.org/10.1016/j.nurpra.2021.02.010; | COVID-19 is a deadly global pandemic with scientific efforts improving our understanding of this novel coronavirus. No proven disease-specific therapies exist although various antiviral regimens offer some success. Many vaccines are in development and phase III clinical trial testing. COVID-19 thrives on medically fragile, elderly and socially disadvantaged, while children have been less affected. Children at risk are those with co-morbidities and neonates; the |

| Title | Authors | Date | Journal | URL | Abstract |
|---|---|---|---|---|---|
| | Alcindor, Magaline A.; Allen, Deborah | | | https://www.ncbi.nlm.nih.gov/pubmed/33723483/; https://www.sciencedirect.com/science/article/pii/S1555415521000660?v=s5 | multisystem inflammatory syndrome is a severe version diagnosed in high-risk children. This article provides COVID-19 management for children, and implications for nursing and advanced practice providers. Suggested Reviewers: Response to Reviewers: As the review was a lot. Please see all of the revisions requested by the reviewers as an attachment in |
| The impact of the COVID-19 pandemic on maternal and perinatal health: a scoping review | Kotlar, Bethany; Gerson, Emily; Petrillo, Sophia; Langer, Ana; Tiemeier, Henning | 2021-01-18 | Reprod Health | https://www.ncbi.nlm.nih.gov/pubmed/33461593/; https://doi.org/10.1186/s12978-021-01070-6 | INTRODUCTION: The Covid-19 pandemic affects maternal health both directly and indirectly, and direct and indirect effects are intertwined. To provide a comprehensive overview on this broad topic in a rapid format behooving an emergent pandemic we conducted a scoping review. METHODS: A scoping review was conducted to compile evidence on direct and indirect impacts of the pandemic on maternal health and provide an overview of the most significant outcomes thus far. Working papers and news articles were considered appropriate evidence along with peer-reviewed publications in order to capture rapidly evolving updates. Literature in English published from January 1st to September 11 2020 was included if it pertained to the direct or indirect effects of the COVID-19 pandemic on the physical, mental, economic, or social health and wellbeing of pregnant people. Narrative descriptions were written about subject areas for which the authors found the most evidence. RESULTS: The search yielded 396 publications, of which 95 were included. Pregnant individuals were found to be at a heightened risk of more severe symptoms than people who are not pregnant. Intrauterine, vertical, and breastmilk transmission were unlikely. Labor, delivery, and breastfeeding guidelines for COVID-19 positive patients varied. Severe increases in maternal mental health issues, such as clinically relevant anxiety and depression, were reported. Domestic violence appeared to spike. Prenatal care visits decreased, healthcare infrastructure was strained, and potentially harmful policies implemented with little evidence. Women were more likely to lose their income due to the pandemic than men, and working mothers struggled with increased childcare demands. CONCLUSION: Pregnant women |

| Title | Authors | Date | Journal | URL | Abstract |
|---|---|---|---|---|---|
| | | | | | and mothers were not found to be at higher risk for COVID-19 infection than people who are not pregnant, however pregnant people with symptomatic COVID-19 may experience more adverse outcomes compared to non-pregnant people and seem to face disproportionate adverse socio-economic consequences. High income and low- and middle-income countries alike faced significant struggles. Further resources should be directed towards quality epidemiological studies. PLAIN ENGLISH SUMMARY: The Covid-19 pandemic impacts reproductive and perinatal health both directly through infection itself but also indirectly as a consequence of changes in health care, social policy, or social and economic circumstances. The direct and indirect consequences of COVID-19 on maternal health are intertwined. To provide a comprehensive overview on this broad topic we conducted a scoping review. Pregnant women who have symptomatic COVID-19 may experience more severe outcomes than people who are not pregnant. Intrauterine and breastmilk transmission, and the passage of the virus from mother to baby during delivery are unlikely. The guidelines for labor, delivery, and breastfeeding for COVID-19 positive patients vary, and this variability could create uncertainty and unnecessary harm. Prenatal care visits decreased, healthcare infrastructure was strained, and potentially harmful policies are implemented with little evidence in high and low/middle income countries. The social and economic impact of COVID-19 on maternal health is marked. A high frequency of maternal mental health problems, such as clinically relevant anxiety and depression, during the epidemic are reported in many countries. This likely reflects an increase in problems, but studies demonstrating a true change are lacking. Domestic violence appeared to spike. Women were more vulnerable to losing their income due to the pandemic than men, and working mothers struggled with increased childcare demands. We make several recommendations: more resources should be directed to epidemiological studies, health and social services for pregnant women and mothers should not be diminished, and more focus on maternal mental health during the epidemic is needed. |

| Title | Authors | Date | Journal | URL | Abstract |
| --- | --- | --- | --- | --- | --- |
| Effect of SARS-CoV-2 Infection in Pregnancy on Maternal and Neonatal Outcomes in Africa: An AFREhealth Call for Evidence through Multicountry Research Collaboration | Nachega, Jean B.; Sam-Agudu, Nadia A.; Budhram, Samantha; Taha, Taha E.; Vannevel, Valerie; Somapillay, Priya; Ishoso, Daniel Katuashi; Tshiasuma Pipo, Michel; Bongo-Pasi Nswe, Christian; Ditekemena, John; Ayele, Birhanu T.; Machekano, Rhoderick N.; Gachuno, Onesmus W.; Kinuthia, John; Mwongeli, Nancy; Sekikubo, Musa; Musoke, Philippa; Agbeno, Evans Kofi; Umar, Lawal W.; Ntakwinja, Mukanire; Mukwege, Denis M.; Smith, Emily R.; Mills, Eduard J.; Otshudiema, John Otokoye; Mbala-Kingebeni, Placide; Kayembe, Jean-Marie N.; Mavungu Landu, Don Jethro; Muyembe Tamfum, Jean-Jacques; Zumla, Alimuddin; | 2020-12-28 | Am J Trop Med Hyg | https://doi.org/10.4269/ajtmh.20-1553; https://www.ncbi.nlm.nih.gov/pubmed/33372651/ | In the African context, there is a paucity of data on SARS-CoV-2 infection and associated COVID-19 in pregnancy. Given the endemicity of infections such as malaria, HIV, and tuberculosis (TB) in sub-Saharan Africa (SSA), it is important to evaluate coinfections with SARS-CoV-2 and their impact on maternal/infant outcomes. Robust research is critically needed to evaluate the effects of the added burden of COVID-19 in pregnancy, to help develop evidence-based policies toward improving maternal and infant outcomes. In this perspective, we briefly review current knowledge on the clinical features of COVID-19 in pregnancy; the risks of preterm birth and cesarean delivery secondary to comorbid severity; the effects of maternal SARS-CoV-2 infection on the fetus/neonate; and in utero mother-to-child SARS-CoV-2 transmission. We further highlight the need to conduct multicountry surveillance as well as retrospective and prospective cohort studies across SSA. This will enable assessments of SARS-CoV-2 burden among pregnant African women and improve the understanding of the spectrum of COVID-19 manifestations in this population, which may be living with or without HIV, TB, and/or other coinfections/comorbidities. In addition, multicountry studies will allow a better understanding of risk factors and outcomes to be compared across countries and subregions. Such an approach will encourage and strengthen much-needed intra-African, south-to-south multidisciplinary and interprofessional research collaborations. The African Forum for Research and Education in Health's COVID-19 Research Working Group has embarked upon such a collaboration across Western, Central, Eastern and Southern Africa. |

| Title | Authors | Date | Journal | URL | Abstract |
|---|---|---|---|---|---|
| | Langenegger, Eduard J.; Mofenson, Lynne M. | | | | |
| COVID-19 in Malawi: lessons in pandemic preparedness from a tertiary children's hospital | Chaziya, Jessica; Freyne, Bridget; Lissauer, Samantha; Nielsen, Maryke; Langton, Josephine; O'Hare, Bernadette; Molyneux, Liz; Moxon, Christopher; Iroh Tam, Pui-Ying; Hoskyns, Lucy; Masanjala, Henderson; Ilepere, Sakina; Ngwira, Memory; Kawaza, Kondwani; Mumba, Daniel; Chimalizeni, Yamikani; Dube, Queen | 2020-12-23 | Arch Dis Child | https://doi.org/10.1136/archdischild-2020-319980; https://www.ncbi.nlm.nih.gov/pubmed/33361067/ | |
| COVID-19 PICU guidelines: for high- and limited-resource settings | Kache, Saraswati; Chisti, Mohammod Jobayer; Gumbo, Felicity; Mupere, Ezekiel; Zhi, Xia; Nallasamy, Karthi; Nakagawa, Satoshi; Lee, Jan Hau; Di Nardo, Matteo; de la Oliva, Pedro; Katyal, Chhavi; Anand, Kanwaljeet J. S.; de Souza, Daniela Carla; | 2020-07-07 | Pediatr Res | https://www.ncbi.nlm.nih.gov/pubmed/32634818/; https://doi.org/10.1038/s41390-020-1053-9 | BACKGROUND: Fewer children than adults have been affected by the COVID-19 pandemic, and the clinical manifestations are distinct from those of adults. Some children particularly those with acute or chronic co-morbidities are likely to develop critical illness. Recently, a multisystem inflammatory syndrome (MIS-C) has been described in children with some of these patients requiring care in the pediatric ICU. METHODS: An international collaboration was formed to review the available evidence and develop evidence-based guidelines for the care of critically ill children with SARS-CoV-2 infection. Where the evidence was lacking, those gaps were replaced with consensus-based guidelines. RESULTS: This process has generated 44 recommendations related to pediatric COVID-19 patients presenting with respiratory distress or failure, sepsis or septic shock, cardiopulmonary arrest, MIS-C, those requiring adjuvant |

| Title | Authors | Date | Journal | URL | Abstract |
|---|---|---|---|---|---|
| | Lanziotti, Vanessa Soares; Carcillo, Joseph | | | | therapies, or ECMO. Evidence to explain the milder disease patterns in children and the potential to use repurposed anti-viral drugs, anti-inflammatory or anti-thrombotic therapies are also described. CONCLUSION: Brief summaries of pediatric SARS-CoV-2 infection in different regions of the world are included since few registries are capturing this data globally. These guidelines seek to harmonize the standards and strategies for intensive care that critically ill children with COVID-19 receive across the world. IMPACT: At the time of publication, this is the latest evidence for managing critically ill children infected with SARS-CoV-2. Referring to these guidelines can decrease the morbidity and potentially the mortality of children effected by COVID-19 and its sequalae. These guidelines can be adapted to both high- and limited-resource settings. |
| A Case for Girl-child Education to Prevent and Curb the Impact of Emerging Infectious Diseases Epidemics | Frimpong, Shadrack; Paintsil, Elijah | 2020-09-30 | Yale J Biol Med | https://www.ncbi.nlm.nih.gov/pmc/articles/PMC7513442/ | Not only do epidemics such as HIV/AIDS, Ebola Virus Disease (EVD), and the current Coronavirus Disease (COVID-19) cause the loss of millions of lives, but they also cost the global economy billions of dollars. Consequently, there is an urgent need to formulate interventions that will help control their spread and impact when they emerge. The education of young girls and women is one such historical approach. They are usually the vulnerable targets of disease outbreaks – they are most likely to be vehicles for the spread of epidemics due to their assigned traditional roles in resource-limited countries. Based on our work and the work of others on educational interventions, we propose six critical components of a cost-effective and sustainable response to promote girl-child education in resource-limited settings. |
| Surveillance of respiratory viruses among children attending a primary school in | Adema, Irene Wangwa; Kamau, Everlyn; Uchi Nyiro, Joyce; Otieno, Grieven P.; Lewa, Clement; Munywoki, Patrick K.; Nokes, D. James | 2020-09-24 | Wellcome Open Res | https://www.ncbi.nlm.nih.gov/pubmed/33102784/; https://doi.org/10.12688/wellcomeopenres.15703.2 | Background: Respiratory viruses are primary agents of respiratory tract diseases. Knowledge on the types and frequency of respiratory viruses affecting school-children is important in determining the role of schools in transmission in the community and identifying targets for interventions. Methods: We conducted a one-year (term-time) surveillance of respiratory viruses in a rural primary school in Kilifi County, coastal Kenya between May 2017 and April 2018. A sample of 60 students with symptoms of ARI were targeted for nasopharyngeal swab (NPS) collection weekly. Swabs were screened for 15 respiratory virus targets using |

| Title | Authors | Date | Journal | URL | Abstract |
|---|---|---|---|---|---|
| rural coastal Kenya | | | | | real time PCR diagnostics. Data from respiratory virus surveillance at the local primary healthcare facility was used for comparison. Results: Overall, 469 students aged 2-19 years were followed up for 220 days. A total of 1726 samples were collected from 325 symptomatic students; median age of 7 years (IQR 5-11). At least one virus target was detected in 384 (22%) of the samples with a frequency of 288 (16.7%) for rhinovirus, 47 (2.7%) parainfluenza virus, 35 (2.0%) coronavirus, 15 (0.9%) adenovirus, 11 (0.6%) respiratory syncytial virus (RSV) and 5 (0.3%) influenza virus. The proportion of virus positive samples was higher among lower grades compared to upper grades (25.9% vs 17.5% respectively; $\chi$ (2) = 17.2, P -value <0.001). Individual virus target frequencies did not differ by age, sex, grade, school term or class size. Rhinovirus was predominant in both the school and outpatient setting. Conclusion: Multiple respiratory viruses circulated in this rural school population. Rhinovirus was dominant in both the school and outpatient setting and RSV was of notably low frequency in the school. The role of school children in transmitting viruses to the household setting is still unclear and further studies linking molecular data to contact patterns between the school children and their households are required. |
| Tackling Childhood Stunting in the Eastern Mediterranean Region in the Context of COVID-19 | Jawaldeh, Ayoub Al; Doggui, Radhouene; Borghi, Elaine; Aguenaou, Hassan; Ammari, Laila El; Abul-Fadl, Azza; McColl, Karen | 2020-11-19 | Children (Basel) | https://www.ncbi.nlm.nih.gov/pubmed/33227997/; https://doi.org/10.3390/children7110239 | Over 20 million children under 5 years old in the WHO Eastern Mediterranean Region have stunted growth, as a result of chronic malnutrition, with damaging long-term consequences for individuals and societies. This review extracted and analyzed data from the UNICEF, WHO and the World Bank malnutrition estimates to present an overall picture of childhood stunting in the region. The number of children under 5 in the region who are affected by stunting has dropped from 24.5 million (40%) in 1990 to 20.6 million (24.2%) in 2019. The reduction rate since the 2012 baseline is only about two fifths of that required and much more rapid progress will be needed to reach the internationally agreed targets by 2025 and 2030. Prevalence is highest in low-income countries and those with a lower Human Development Index. The COVID-19 pandemic threatens to undermine efforts to reduce stunting, through its impact on access and affordability of safe and nutritious foods and access to important health |

| Title | Authors | Date | Journal | URL | Abstract |
|---|---|---|---|---|---|
| | | | | | services. Priority areas for action to tackle stunting as part of a comprehensive, multisectoral nutrition strategy are proposed. In light of the threat that COVID-19 will exacerbate the already heavy burden of malnutrition in the Eastern Mediterranean Region, implementation of such strategies is more important than ever. |
| The Burden of Malnutrition and Fatal COVID-19: A Global Burden of Disease Analysis | Mertens, Elly; Peñalvo, José L. | 2021-01-21 | Front Nutr | https://doi.org/10.3389/fnut.2020.619850; https://www.ncbi.nlm.nih.gov/pubmed/33553234/ | Background: Although reasonable to assume, it is not yet clear whether malnourished countries are at higher risk for severe or fatal coronavirus disease 2019 (COVID-19). This study aims to identify the countries where prevalent malnutrition may be a driving factor for fatal disease after severe acute respiratory syndrome coronavirus 2 (SARS-CoV-2) infection. Methods: Using estimates from the Global Burden of Disease 2019, country-level burden of malnutrition was quantified using four indicators: death rates for child growth failure (underweight, stunting, and/or wasting) and years lived with disability (YLD) attributed to iron and vitamin A deficiencies and high body mass index (BMI). Global mortality descriptors of the ongoing COVID-19 pandemic were extracted from the European Centre for Disease Prevention and Control, and case fatality ratios (CFRs) were calculated introducing a lag time of 10 weeks after the first death of a confirmed case. Bivariate analyses for 172 countries were carried out for malnutrition indicators and fatal COVID-19. Correlations between burden indicators were characterized by Spearman's rank correlation coefficients (ρ) and visually by scatterplots. Restricted cubic splines and underlying negative binomial regressions adjusted for countries' age-structure, prevalent chronic comorbidities related to COVID-19, population density, and income group were used to explore non-linear relationships. Results: Stratified by the World Bank income group, a moderate positive association between YLD rates for iron deficiency and CFRs for COVID-19 was observed for low-income countries ($\rho = 0.60$, $p = 0.027$), whereas no clear indications for the association with child growth failure, vitamin A deficiency, or high BMI were found ($\rho < 0.30$). Countries ranking high on at least three malnutrition indicators and presenting also an elevated CFR for COVID-19 are sub-Saharan African countries, namely, |

| Title | Authors | Date | Journal | URL | Abstract |
|---|---|---|---|---|---|
| | | | | | Angola, Burkina Faso, Chad, Liberia, Mali, Niger, Sudan, and Tanzania, as well as Yemen and Guyana. Conclusions: Population-level malnutrition appears to be related to increased rates of fatal COVID-19 in areas with an elevated burden of undernutrition, such as countries in the Sahel strip. COVID-19 response plans in malnourished countries, vulnerable to fatal COVID-19, should incorporate food security, nutrition, and social protection as a priority component in order to reduce COVID-19 fatality. |
| Health systems and nutrition in the time of COVID-19 | Khan, Amir Ullah; Bali, Vinita | 2021-03-18 | J | https://www.ncbi.nlm.nih.gov/pmc/articles/PMC7971388/ | As infection rates rise, job losses increase and workers leave cities to walk back home, and there is a silent hunger and nutrition crisis striking the country. Those who will bear the brunt of this are the already vulnerable—namely, children, adolescent girls, nursing and expectant mothers—now denied even basic calories. Among these are some who are also suffering huge weight losses because of the 15 days of high fever. This tragedy will play out in various horrifying ways in the future and must be addressed with urgency. Our stimulus package promises loans, which will take time to reach the poor, and a meager ration of cereals and pulses, while hunger and insufficient nutrition are immediate problems as Raghuram Rajan pointed out recently. |
| Early estimates of the indirect effects of the COVID-19 pandemic on maternal and child mortality in low-income and middle-income countries: a | Roberton, Timothy; Carter, Emily D; Chou, Victoria B; Stegmuller, Angela R; Jackson, Bianca D; Tam, Yvonne; Sawadogo-Lewis, Talata; Walker, Neff | 2020-05-12 | Lancet Glob Health | https://api.elsevier.com/content/article/pii/S2214109X20302291; https://www.sciencedirect.com/science/article/pii/S2214109X20302291; https://doi.org/10.1016/s2214-109x(20)30229-1; https://www.ncbi.nlm.nih.gov/pubmed/32405459/ | BACKGROUND: While the COVID-19 pandemic will increase mortality due to the virus, it is also likely to increase mortality indirectly. In this study, we estimate the additional maternal and under-5 child deaths resulting from the potential disruption of health systems and decreased access to food. METHODS: We modelled three scenarios in which the coverage of essential maternal and child health interventions is reduced by 9·8–51·9% and the prevalence of wasting is increased by 10–50%. Although our scenarios are hypothetical, we sought to reflect real-world possibilities, given emerging reports of the supply-side and demand-side effects of the pandemic. We used the Lives Saved Tool to estimate the additional maternal and under-5 child deaths under each scenario, in 118 low-income and middle-income countries. We estimated additional deaths for a single month and extrapolated for 3 months, 6 months, and 12 months. FINDINGS: Our least severe scenario (coverage reductions of 9·8–18·5% and wasting increase of |

| Title | Authors | Date | Journal | URL | Abstract |
|---|---|---|---|---|---|
| modelling study | | | | | 10%) over 6 months would result in 253 500 additional child deaths and 12 200 additional maternal deaths. Our most severe scenario (coverage reductions of 39·3–51·9% and wasting increase of 50%) over 6 months would result in 1 157 000 additional child deaths and 56 700 additional maternal deaths. These additional deaths would represent an increase of 9·8–44·7% in under-5 child deaths per month, and an 8·3–38·6% increase in maternal deaths per month, across the 118 countries. Across our three scenarios, the reduced coverage of four childbirth interventions (parenteral administration of uterotonics, antibiotics, and anticonvulsants, and clean birth environments) would account for approximately 60% of additional maternal deaths. The increase in wasting prevalence would account for 18–23% of additional child deaths and reduced coverage of antibiotics for pneumonia and neonatal sepsis and of oral rehydration solution for diarrhoea would together account for around 41% of additional child deaths. INTERPRETATION: Our estimates are based on tentative assumptions and represent a wide range of outcomes. Nonetheless, they show that, if routine health care is disrupted and access to food is decreased (as a result of unavoidable shocks, health system collapse, or intentional choices made in responding to the pandemic), the increase in child and maternal deaths will be devastating. We hope these numbers add context as policy makers establish guidelines and allocate resources in the days and months to come. FUNDING: Bill & Melinda Gates Foundation, Global Affairs Canada. |
| Possible Impact of COVID-19 on Children in Africa, Reflections from Italy and Burkina Faso | Ouedraogo, Paul; Schumacher, Richard Fabian | 2020-08-07 | J Trop Pediatr | https://www.ncbi.nlm.nih.gov/pubmed/32766698/; https://doi.org/10.1093/tropej/fmaa055 | Africa is the World Health Organization-region least affected by the Severe Acute Respiratory Syndrome Coronavirus 2 (SARS-CoV-2) pandemic. Here, we compare the situation in severely hit Italy with that in less hit Burkina Faso, focussing on the differences in epidemiological, geographical, demographical, cultural and medical conditions to highlight how a full-blown war on the pandemic can impact on other, equally important aspects of global child health. |

| Title | Authors | Date | Journal | URL | Abstract |
|---|---|---|---|---|---|
| Food availability, accessibility and dietary practices during the COVID-19 pandemic: a multi-country survey | Jafri, Ali; Mathe, Nonsikelelo; Aglago, Elom K; Konyole, Silvenus O; Ouedraogo, Moussa; Audain, Keiron; Zongo, Urbain; Laar, Amos K; Johnson, Jeffrey; Sanou, Dia | 2021-03-05 | Public health nutrition | https://www.ncbi.nlm.nih.gov/pubmed/33663623/; https://doi.org/10.1017/s1368980021000987 | OBJECTIVE: To investigate the perceived effects of the coronavirus disease (COVID-19) pandemic lockdown measures on food availability, accessibility, dietary practices and strategies used by participants to cope with these measures. DESIGN: We conducted a cross-sectional multi-country online survey between May and July 2020. We used a study-specific questionnaire mainly based on the adaptation of questions to assess food security and coping strategies from the World Food Programme's 'Emergency Food Security Assessment' and 'The Coping Strategy Index'. SETTING: The questionnaire was hosted online using Google Forms and shared using social media platforms. PARTICIPANTS: A total of 1075 adult participants from eighty-two countries completed the questionnaire. RESULTS: As a prelude to COVID-19 lockdowns, 62·7 % of the participants reported to have stockpiled food, mainly cereals (59·5 % of the respondents) and legumes (48·8 %). An increase in the prices of staples, such as cereals and legumes, was widely reported. Price increases have been identified as an obstacle to food acquisition by 32·7 % of participants. Participants reported having lesser variety (50·4 %), quality (30·2 %) and quantity (39·2 %) of foods, with disparities across regions. Vulnerable groups were reported to be facing some struggle to acquire adequate food, especially people with chronic diseases (20·2 %), the elderly (17·3 %) and children (14·5 %). To cope with the situation, participants mostly relied on less preferred foods (49 %), reduced portion sizes (30 %) and/or reduced the number of meals (25·7 %). CONCLUSIONS: The COVID-19 pandemic negatively impacted food accessibility and availability, altered dietary practices and worsened the food insecurity situation, particularly in the most fragile regions. |
| Predicting the Impact of COVID-19 and the Potential Impact of the | Bell, David; Hansen, Kristian Schultz; Kiragga, Agnes N.; Kambugu, Andrew; Kissa, John; Mbonye, Anthony K. | 2020-07-23 | Am J Trop Med Hyg | https://www.ncbi.nlm.nih.gov/pubmed/32705975/; https://doi.org/10.4269/ajtmh.20-0546 | The COVID-19 pandemic and public health "lockdown" responses in sub-Saharan Africa, including Uganda, are now widely reported. Although the impact of COVID-19 on African populations has been relatively light, it is feared that redirecting focus and prioritization of health systems to fight COVID-19 may have an impact on access to non–COVID-19 diseases. We applied age-based COVID-19 mortality data from China to the population structures of Uganda and |

| Title | Authors | Date | Journal | URL | Abstract |
|---|---|---|---|---|---|
| Public Health Response on Disease Burden in Uganda | | | | | non-African countries with previously established outbreaks, comparing theoretical mortality and disability-adjusted life years (DALYs) lost. We then predicted the impact of possible scenarios of the COVID-19 public health response on morbidity and mortality for HIV/AIDS, malaria, and maternal health in Uganda. Based on population age structure alone, Uganda is predicted to have a relatively low COVID-19 burden compared with an equivalent transmission in comparison countries, with 12% of the mortality and 19% of the lost DALYs predicted for an equivalent transmission in Italy. By contrast, scenarios of the impact of the public health response on malaria and HIV/AIDS predict additional disease burdens outweighing that predicted from extensive SARS-CoV-2 transmission. Emerging disease data from Uganda suggest that such deterioration may already be occurring. The results predict a relatively low COVID-19 impact on Uganda associated with its young population, with a high risk of negative impact on non–COVID-19 disease burden from a prolonged lockdown response. This may reverse hard-won gains in addressing fundamental vulnerabilities in women and children's health, and underlines the importance of tailoring COVID-19 responses according to population structure and local disease vulnerabilities. |
| The Changing Aspects of Motherhood in Face of the COVID-19 Pandemic in Low- and Middle-Income Countries | Kingsley, Jennifer Prince; Vijay, Paul Kingsley; Kumaresan, Jacob; Sathiakumar, Nalini | 2020-11-26 | Matern Child Health J | https://doi.org/10.1007/s10995-020-03044-9; https://www.ncbi.nlm.nih.gov/pubmed/33244678/ | PURPOSE: To advocate perspectives to strengthen existing healthcare systems to prioritize maternal health services amidst and beyond the COVID-19 pandemic in low- and middle income countries. DESCRIPTION: COVID-19 directly affects pregnant women causing more severe disease and adverse pregnancy outcomes. The indirect effects due to the monumental COVID-19 response are much worse, increasing maternal and neonatal mortality. ASSESSMENT: Amidst COVID-19, governments must balance effective COVID-19 response measures while continuing delivery of essential health services. Using the World Health Organization's operational guidelines as a base, countries must conduct contextualized analyses to tailor their operations. Evidence based information on different services and comparative cost-benefits will help decisions on trade-offs. Situational analyses identifying extent and reasons for service disruptions and estimates of impacts using modelling |

| Title | Authors | Date | Journal | URL | Abstract |
|---|---|---|---|---|---|
| | | | | | techniques will guide prioritization of services. Ensuring adequate supplies, maintaining core interventions, expanding non-physician workforce and deploying telehealth are some adaptive measures to optimize care. Beyond the COVID-19 pandemic, governments must reinvest in maternal and child health by building more resilient maternal health services supported by political commitment and multisectoral engagement, and with assistance from international partners. CONCLUSIONS: Multi-sectoral investments providing high-quality care that ensures continuity and available to all segments of the population are needed. A robust primary healthcare system linked to specialist care and accessible to all segments of the population including marginalized subgroups is of paramount importance. Systematic approaches to digital health care solutions to bridge gaps in service is imperative. Future pandemic preparedness programs must include action plans for resilient maternal health services. |
| Beyond the Disease: Contextualized Implications of the COVID-19 Pandemic for Children and Young People Living in Eastern and Southern Africa | Govender, Kaymarlin; Cowden, Richard Gregory; Nyamaruze, Patrick; Armstrong, Russell Murray; Hatane, Luann | 2020-10-19 | Front Public Health | https://doi.org/10.3389/fpubh.2020.00504; https://www.ncbi.nlm.nih.gov/pubmed/33194933/ | The coronavirus disease 2019 (COVID-19) pandemic has created extraordinary challenges and prompted remarkable social changes around the world. The effects of COVID-19 and the public health control measures that have been implemented to mitigate its impact are likely to be accompanied by a unique set of consequences for specific subpopulations living in low-income countries that have fragile health systems and pervasive social-structural vulnerabilities. This paper discusses the implications of COVID-19 and related public health interventions for children and young people living in Eastern and Southern Africa. Actionable prevention, care, and health promotion initiatives are proposed to attenuate the negative effects of the pandemic and government-enforced movement restrictions on children and young people. |

| Title | Authors | Date | Journal | URL | Abstract |
|---|---|---|---|---|---|
| Falling living standards during the COVID-19 crisis: Quantitative evidence from nine developing countries | Egger, Dennis; Miguel, Edward; Warren, Shana S.; Shenoy, Ashish; Collins, Elliott; Karlan, Dean; Parkerson, Doug; Mobarak, A. Mushfiq; Fink, Günther; Udry, Christopher; Walker, Michael; Haushofer, Johannes; Larreboure, Magdalena; Athey, Susan; Lopez-Pena, Paula; Benhachmi, Salim; Humphreys, Macartan; Lowe, Layna; Meriggi, Niccoló F.; Wabwire, Andrew; Davis, C. Austin; Pape, Utz Johann; Graff, Tilman; Voors, Maarten; Nekesa, Carolyn; Vernot, Corey | 2021-02-05 | Sci Adv | https://doi.org/10.1126/sciadv.abe0997; https://www.ncbi.nlm.nih.gov/pubmed/33547077/ | Despite numerous journalistic accounts, systematic quantitative evidence on economic conditions during the ongoing COVID-19 pandemic remains scarce for most low- and middle-income countries, partly due to limitations of official economic statistics in environments with large informal sectors and subsistence agriculture. We assemble evidence from over 30,000 respondents in 16 original household surveys from nine countries in Africa (Burkina Faso, Ghana, Kenya, Rwanda, Sierra Leone), Asia (Bangladesh, Nepal, Philippines), and Latin America (Colombia). We document declines in employment and income in all settings beginning March 2020. The share of households experiencing an income drop ranges from 8 to 87% (median, 68%). Household coping strategies and government assistance were insufficient to sustain precrisis living standards, resulting in widespread food insecurity and dire economic conditions even 3 months into the crisis. We discuss promising policy responses and speculate about the risk of persistent adverse effects, especially among children and other vulnerable groups. |
| Syndemic effects in complex humanitarian emergencies: a framework for understandin | Brandon A, Kohrt; Lauren, Carruth | 2020-09-19 | Soc Sci Med | https://www.sciencedirect.com/science/article/pii/S0277953620305979?v=s5; https://www.ncbi.nlm.nih.gov/pubmed/33051023/; https://doi.org/10.101 | A hallmark of complex humanitarian emergencies is the collective exposure, often over extended periods of time, to political violence in the forms of war, terrorism, political intimidation, repression, unlawful detention, and forced displacement. Populations in complex humanitarian emergencies have higher risks of multiple co-morbidities: mental disorders, infectious diseases, malnutrition, and chronic non-communicable physical diseases. However, there is wide variation in the health impacts of both across and within humanitarian emergencies. Syndemic theory is an approach to conceptualizing multiple |

| Title | Authors | Date | Journal | URL | Abstract |
|---|---|---|---|---|---|
| g political violence and improving multi-morbidity health outcomes | | | | 6/j.socscimed.2020.113378; https://api.elsevier.com/content/article/pii/S0277953620305979 | morbidities and social determinants to understand differential patterns of multimorbidity, elucidate underlying mechanisms, and better design interventions. Syndemic theory, if applied to complex humanitarian emergencies, has the potential to uncover origins of localized patterns of multi-morbidity resulting from political violence and historical inequities. In this paper, we present two case studies based on mixed-methods research to illustrate how syndemic models can be applied in complex humanitarian emergencies. First, in a Nepal case study, we explore different patterns of posttraumatic stress disorder (PTSD) and depression co-morbidity among female former child soldiers returning home after war. Despite comparable exposure to war-related traumas, girl soldiers in high-caste Hindu communities had 63% co-morbidity of PTSD and depression, whereas girl soldiers in communities with mixed castes and religions, had 8% PTSD prevalence, but no cases of PTSD and depression co-morbidity. In the second case study, we explore the high rates of type 2 diabetes during a spike in political violence and population displacement. Despite low rates of obesity and other common risk factors, Somalis in Ethiopia experienced rising cases of and poor outcomes from type-2 diabetes. Political violence shapes healthcare resources, diets, and potentially, this epidemiological anomaly. Based on these case studies we propose a humanitarian syndemic research agenda for observational and intervention studies, with the central focus being that public health efforts need to target violence prevention at family, community, national, and global levels. |
| Child Maltreatment in the Time of the COVID-19 Pandemic: A Proposed Global | Katz, Carmit; Priolo Filho, Sidnei R.; Korbin, Jill; Bérubé, Annie; Fouché, Ansie; Haffejee, Sadiyya; Kaawa-Mafigiri, David; Maguire-Jack, Kathryn; Muñoz, Pablo; | 2020-11-20 | Child Abuse Negl | https://api.elsevier.com/content/article/pii/S0145213420304798; https://doi.org/10.1016/j.chiabu.2020.104824; https://www.sciencedirect.com/science/articl | BACKGROUND: Child protection is and will be drastically impacted by the COVID-19 pandemic. Comprehending this new reality and identifying research, practice and policy paths are urgent needs. OBJECTIVE: The current paper aims to suggest a framework for risk and protective factors that need to be considered in child protection in its various domains of research, policy, and practice during and after the COVID-19 pandemic. STRATEGY: From an international collaboration involving researchers and child protection professionals from eight countries, the current paper examines various factors that were identified as |

| Title | Authors | Date | Journal | URL | Abstract |
|---|---|---|---|---|---|
| Framework on Research, Policy and Practice | Spilsbury, James; Tarabulsy, George; Tiwari, Ashwini; Levine Thembekile, Diane; Truter, Elmein; Varela, Natalia | | | e/pii/S0145213420304798?v=s5; https://www.ncbi.nlm.nih.gov/pubmed/33353782/ | playing an important role in the child protection system. THE INITIAL SUGGESTED FRAMEWORK: Through the use of an ecological framework, the current paper points to risk and protective factors that need further exploration. Key conclusions point to the urgent need to address the protection of children in this time of a worldwide pandemic. Discussion of risk and protective factors is significantly influenced by the societal context of various countries, which emphasizes the importance of international collaboration in protecting children, especially in the time of a worldwide pandemic. CONCLUSION: The COVID-19 pandemic has stressed the urgent need to advance both theory and practice in order to ensure children's rights to safety and security during any pandemic. The suggested framework has the potential to advance these efforts so that children will be better protected from maltreatment amidst a pandemic in the future. |
| Spillover Trends of Child Labor During the Coronavirus Crisis- an Unnoticed Wake-Up Call | Ahad, Md. Abdul; Parry, Yvonne K.; Willis, Eileen | 2020-09-04 | Front Public Health | https://www.ncbi.nlm.nih.gov/pubmed/33014979/; https://doi.org/10.3389/fpubh.2020.00488 | |
| Is Nigeria prepared and ready to respond to the COVID-19 pandemic in its conflict- | Tijjani, Salman Jidda; Ma, Le | 2020-05-27 | Int J Equity Health | https://www.ncbi.nlm.nih.gov/pubmed/32460766/; https://doi.org/10.1186/s12939-020-01192-6 | Northeastern Nigeria has over the decade suffered from the Boko Haram insurgency and is still in the process of recovery from the complex humanitarian crisis that has displaced and subjected millions of vulnerable children, women and elderly population to poverty, disease outbreaks, hunger and malnutrition. Yet, the conflict-affected states in Northeastern Nigeria is not far away from being the worse-hit by the COVID-19 pandemic if urgent public health preventive measures are not taken to contain the spread of the deadly and highly |

| Title | Authors | Date | Journal | URL | Abstract |
| --- | --- | --- | --- | --- | --- |
| affected northeastern states? | | | | | infectious virus. The question arises, “what is Nigeria doing to tackle the burden of a COVID-19 spread and an ongoing humanitarian crisis? |
| The potential impact of the COVID-19 pandemic on child growth and development: a systematic review() | Araújo, Liubiana Arantes de; Veloso, Cássio Frederico; Souza, Matheus de Campos; Azevedo, João Marcos Coelho de; Tarro, Giulio | 2020-09-23 | J Pediatr (Rio J) | https://www.sciencedirect.com/science/article/pii/S0021755720302096?v=s5; https://www.ncbi.nlm.nih.gov/pubmed/32980318/; https://doi.org/10.1016/j.jped.2020.08.008; https://api.elsevier.com/content/article/pii/S0021755720302096 | OBJECTIVE: This was a systematic review of studies that examined the impact of epidemics or social restriction on mental and developmental health in parents and children/adolescents. SOURCE OF DATA: The PubMed, WHO COVID-19, and SciELO databases were searched on March 15, 2020, and on April 25, 2020, filtering for children (0–18 years) and humans. SYNTHESIS OF DATA: The tools used to mitigate the threat of a pandemic such as COVID-19 may very well threaten child growth and development. These tools — such as social restrictions, shutdowns, and school closures — contribute to stress in parents and children and can become risk factors that threaten child growth and development and may compromise the Sustainable Development Goals. The studies reviewed suggest that epidemics can lead to high levels of stress in parents and children, which begin with concerns about children becoming infected. These studies describe several potential mental and emotional consequences of epidemics such as COVID-19, H1N1, AIDS, and Ebola: severe anxiety or depression among parents and acute stress disorder, post-traumatic stress, anxiety disorders, and depression among children. These data can be related to adverse childhood experiences and elevated risk of toxic stress. The more adverse experiences, the greater the risk of developmental delays and health problems in adulthood, such as cognitive impairment, substance abuse, depression, and non-communicable diseases. CONCLUSION: Information about the impact of epidemics on parents and children is relevant to policy makers to aid them in developing strategies to help families cope with epidemic/pandemic-driven adversity and ensure their children’s healthy development. |
| Identifying and combating the impacts | Rogerson, Stephen J.; Beeson, James G.; Laman, Moses; Poespoprodjo, Jeanne | 2020-07-30 | BMC Med | https://doi.org/10.1186/s12916-020-01710-x; https://www.ncbi.nlm. | BACKGROUND: The COVID-19 pandemic has resulted in millions of infections, hundreds of thousands of deaths and major societal disruption due to lockdowns and other restrictions introduced to limit disease spread. Relatively little attention has been paid to understanding how the pandemic has affected |

| Title | Authors | Date | Journal | URL | Abstract |
| --- | --- | --- | --- | --- | --- |
| of COVID-19 on malaria | Rini; William, Timothy; Simpson, Julie A.; Price, Ric N. | | | nih.gov/pubmed/32727467/ | treatment, prevention and control of malaria, which is a major cause of death and disease and predominantly affects people in less well-resourced settings. MAIN BODY: Recent successes in malaria control and elimination have reduced the global malaria burden, but these gains are fragile and progress has stalled in the past 5 years. Withdrawing successful interventions often results in rapid malaria resurgence, primarily threatening vulnerable young children and pregnant women. Malaria programmes are being affected in many ways by COVID-19. For prevention of malaria, insecticide-treated nets need regular renewal, but distribution campaigns have been delayed or cancelled. For detection and treatment of malaria, individuals may stop attending health facilities, out of fear of exposure to COVID-19, or because they cannot afford transport, and health care workers require additional resources to protect themselves from COVID-19. Supplies of diagnostics and drugs are being interrupted, which is compounded by production of substandard and falsified medicines and diagnostics. These disruptions are predicted to double the number of young African children dying of malaria in the coming year and may impact efforts to control the spread of drug resistance. Using examples from successful malaria control and elimination campaigns, we propose strategies to re-establish malaria control activities and maintain elimination efforts in the context of the COVID-19 pandemic, which is likely to be a long-term challenge. All sectors of society, including governments, donors, private sector and civil society organisations, have crucial roles to play to prevent malaria resurgence. Sparse resources must be allocated efficiently to ensure integrated health care systems that can sustain control activities against COVID-19 as well as malaria and other priority infectious diseases. CONCLUSION: As we deal with the COVID-19 pandemic, it is crucial that other major killers such as malaria are not ignored. History tells us that if we do, the consequences will be dire, particularly in vulnerable populations. |
| Funding and COVID-19 research | Antonio, Emilia; Alobo, Moses; Tufet Bayona, | 2020-11-24 | AAS Open Res | https://www.ncbi.nlm.nih.gov/pubmed/33709054/; | Background: Emerging data from Africa indicates remarkably low numbers of reported COVID-19 deaths despite high levels of disease transmission. However, evolution of these trends as the pandemic progresses remains unknown. More |

| Title | Authors | Date | Journal | URL | Abstract |
|---|---|---|---|---|---|
| priorities - are the research needs for Africa being met? | Marta; Marsh, Kevin; Norton, Alice | | | https://doi.org/10.12688/aasopenres.13162.1 | certain are the devastating long-term impacts of the pandemic on health and development evident globally. Research tailored to the unique needs of African countries is crucial. UKCDR and GloPID-R have launched a tracker of funded COVID-19 projects mapped to the WHO research priorities and research priorities of Africa and less-resourced countries and published a baseline analysis of a living systematic review (LSR) of these projects. Methods: In-depth analyses of the baseline LSR for COVID-19 funded research projects in Africa (as of 15th July 2020) to determine the funding landscape and alignment of the projects to research priorities of relevance to Africa. Results: The limited COVID-19 related research across Africa appears to be supported mainly by international funding, especially from Europe, although with notably limited funding from United States-based funders. At the time of this analysis no research projects funded by an African-based funder were identified in the tracker although there are several active funding calls geared at research in Africa and there may be funding data that has not been made publicly available. Many projects mapped to the WHO research priorities and five particular gaps in research funding were identified, namely: investigating the role of children in COVID-19 transmission; effective modes of community engagement; health systems research; communication of uncertainties surrounding mother-to-child transmission of COVID-19; and identifying ways to promote international cooperation. Capacity strengthening was identified as a dominant theme in funded research project plans. Conclusions: We found significantly lower funding investments in COVID-19 research in Africa compared to high-income countries, seven months into the pandemic, indicating a paucity of research targeting the research priorities of relevance to Africa. |
| Impact of COVID-19 public health restrictions on older | Giebel, Clarissa; Ivan, Bwire; Burger, Philomena; Ddumba, Isaac | 2020-12-17 | International psychogeriatrics | https://www.ncbi.nlm.nih.gov/pubmed/33327989/; https://doi.org/10.101 | OBJECTIVES: To explore the impact of COVID-19 public health restrictions on the lives of older adults living in Uganda. DESIGN: Qualitative semi-structured interview study. SETTING: Participants' homes. PARTICIPANTS: Older adults living in Uganda (aged 60+). MEASUREMENTS: Older adults in Uganda were interviewed over the phone and asked about their lives before and since COVID- |

| Title | Authors | Date | Journal | URL | Abstract |
|---|---|---|---|---|---|
| people in Uganda: “hunger is really one of those problems brought by this COVID” | | | | 7/s1041610220004081 | 19, and how public health restrictions have affected their lives. Semi-structured interviews were audio-recorded, transcribed and translated into English. Transcripts were thematically analyzed and themes generated in discussion. RESULTS: In total, 30 older adults participated in the study. Five themes were identified: (1) economic impacts; (2) lack of access to basic necessities; (3) impact on healthcare utilization; (4) social impacts and (5) violent reinforcement of public health restrictions. COVID-19 public health restrictions had severe impacts on their lives, with many people having not enough food to eat due to lack of income, and being unable to pay their grandchildren’s school fees. Steep rises in public transport fares and an overall avoidance of transport also resulted in a lack of access to healthcare services and difficulty in getting food. Restrictions were violently reinforced by security guards. CONCLUSIONS: Public health restrictions have a severe impact not only on older adults but also on the whole family in Uganda. Governmental strategies to contain the virus need to provide more support to enable people to get basic necessities and live as normal a life as possible. |
| Food insecurity measurement and prevalence estimates during the COVID-19 pandemic in a repeated cross-sectional survey in Mexico | Gaitán-Rossi, Pablo; Vilar-Compte, Mireya; Teruel, Graciela; Pérez-Escamilla, Rafael | 2020-10-14 | Public health nutrition | https://doi.org/10.1017/s1368980020004000; https://www.ncbi.nlm.nih.gov/pubmed/33050968/ | OBJECTIVE: To validate the telephone modality of the Latin American and Caribbean Food Security Scale (ELCSA) included in three waves of a phone survey to estimate the monthly household food insecurity prevalence during the COVID-19 pandemic in Mexico. DESIGN: We examined the reliability and internal validity of the ELCSA scale in three repeated waves of cross-sectional surveys with Rasch models. We estimated the monthly prevalence of food insecurity in the general population and in households with and without children and compared them with a national 2018 survey. We tested concurrent validity by testing associations of food insecurity with socio-economic status and anxiety. SETTING: ENCOVID-19 is a monthly telephone cross-sectional survey collecting information on the well-being of Mexican households during the pandemic lockdown. Surveys used probabilistic samples, and we used data from April (n 833), May (n 850) and June 2020 (n 1674). PARTICIPANTS: Mexicans 18 years or older who had a mobile telephone. RESULTS: ELCSA had an |

| Title | Authors | Date | Journal | URL | Abstract |
|---|---|---|---|---|---|
| | | | | | adequate model fit and food insecurity was associated, within each wave, with more poverty and anxiety. The COVID-19 lockdown was associated with an important reduction in food security, decreasing stepwise from 38·9 % in 2018 to 24·9 % in June 2020 in households with children. CONCLUSIONS: Telephone surveys were a feasible strategy to monitor reductions in food security during the COVID-19 lockdown. |
| COVID-19 deaths detected in a systematic post-mortem surveillance study in Africa | Mwananyanda, L.; gill, c.; MacLeod, W.; Kwenda, G.; Pieciak, R.; Mupila, Z.; Mupeta, F.; Forman, L.; Ziko, L.; Etter, L.; Thea, D. | 2020-12-24 | | http://medrxiv.org/cgi/content/short/2020.12.22.20248327v1?rss=1; https://doi.org/10.1101/2020.12.22.20248327 | Objectives Limited SARS CoV 2 testing in many African countries has constrained availability of data on the impact of COVID-19 (CV19). To address this gap, we conducted a systematic post-mortem surveillance study to directly measure the fatal impact of CV19 in an urban African population. Design We enrolled deceased individuals at the University Teaching Hospital (UTH) Morgue in Lusaka, Zambia. We obtained nasopharyngeal swabs for testing via reverse-transcriptase quantitative PCR (RT-qPCR) against the SARS-2 Coronavirus. We stratified deaths by CV19 status, by location, age, sex, and underlying risk factors. Setting UTH is the largest tertiary care referral hospital in Zambia and its morgue registers ~80% of Lusaka's deaths. Participants Participants of all ages were enrolled if within 48 hours of death and if the next of kin or representative provided written informed consent. Results We enrolled 372 participants between June and September 2020, and had PCR results for 364 (99.5%). CV19 was detected in 70/364 (19.2%). The median age for CV19+ deaths was 48 years (IQR 36-72 years) and 70% were male. Most CV19+ deaths (51/70, 72.8%) occurred in the community; none had been tested for CV19 antemortem. Among the 19/70 facility deaths, six were tested antemortem. Among the 52/70 CV19 deaths with symptoms data, 44/52 had typical symptoms of CV19 (cough, fever, shortness of breath), of whom only five were tested antemortem. We identified CV19 among seven children; only one had been tested antemortem. The proportion of CV19+ deaths increased with age, but 75.7% of CV19+ deaths were aged <60 years. The five most common co-morbidities among CV19+ deaths were: tuberculosis (31.4%); hypertension (27.1%); HIV/AIDS (22.9%); alcohol use (17.1%); and diabetes (12.9%). |

| Title | Authors | Date | Journal | URL | Abstract |
| --- | --- | --- | --- | --- | --- |
| | | | | | Conclusions Contrary to expectations, CV19+ deaths were common in Lusaka. The majority occurred in the community where testing capacity is lacking. Yet few who died at facilities were tested, despite presenting with typical symptoms of CV19. Therefore, CV19 cases were under reported because testing was rarely done, not because CV19 was rare. If our data are generalizable, the impact of CV19 in Africa has been vastly underestimated. |
| Reorienting Nurturing Care for Early Childhood Development during the COVID-19 Pandemic in Kenya: A Review | Shumba, Constance; Maina, Rose; Mbuthia, Gladys; Kimani, Rachel; Mbugua, Stella; Shah, Sweta; Abubakar, Amina; Luchters, Stanley; Shaibu, Sheila; Ndirangu, Eunice | 2020-09-25 | Int J Environ Res Public Health | https://www.ncbi.nlm.nih.gov/pubmed/32992966/; https://doi.org/10.3390/ijerph17197028 | In Kenya, millions of children have limited access to nurturing care. With the Coronavirus disease 2019 (COVID-19) pandemic, it is anticipated that vulnerable children will bear the biggest brunt of the direct and indirect impacts of the pandemic. This review aimed to deepen understanding of the effects of COVID-19 on nurturing care from conception to four years of age, a period where the care of children is often delivered through caregivers or other informal platforms. The review has drawn upon the empirical evidence from previous pandemics and epidemics, and anecdotal and emerging evidence from the ongoing COVID-19 crisis. Multifactorial impacts fall into five key domains: direct health; health and nutrition systems; economic protection; social and child protection; and child development and early learning. The review proposes program and policy strategies to guide the reorientation of nurturing care, prevent the detrimental effects associated with deteriorating nurturing care environments, and support the optimal development of the youngest and most vulnerable children. These include the provision of cash transfers and essential supplies for vulnerable households and strengthening of community-based platforms for nurturing care. Further research on COVID-19 and the ability of children's ecology to provide nurturing care is needed, as is further testing of new ideas. |
| Demographic, health, and economic transitions and the | King, Elizabeth M.; Randolph, Hannah L.; Floro, Maria S.; Suh, Jooyeoun | 2021-01-18 | World Dev | https://www.sciencedirect.com/science/article/pii/S0305750X2030499X; https://doi.org/10.1016/j.worlddev.2020.10 | The COVID-19 pandemic has caused millions of infections and deaths worldwide, forced schools to suspend classes, workers to work from home, many to lose their livelihoods, and countless businesses to close. Throughout this crisis, families have had to protect, comfort and care for their children, their elderly and other members. While the pandemic has greatly intensified family care responsibilities for families, unpaid care work has been a primary activity of |

| Title | Authors | Date | Journal | URL | Abstract |
|---|---|---|---|---|---|
| future care burden | | | | 5371; https://api.elsevier.com/content/article/pii/S0305750X2030499X; https://www.ncbi.nlm.nih.gov/pubmed/33519035/ | families even in normal times. This paper estimates the future global need for caregiving, and the burden of that need that typically falls on families, especially women. It takes into account projected demographic shifts, health transitions, and economic changes in order to obtain an aggregate picture of the care need relative to the potential supply of caregiving in low-, middle- and high-income countries. This extensive margin of the future care burden, however, does not capture the weight of that burden unless the quantity and quality of care time per caregiver are taken into account. Adjusting for care time given per caregiver, the paper incorporates data from time-use surveys, illustrating this intensive margin of the care burden in three countries that have very different family and economic contexts—Ghana, Mongolia, and South Korea. Time-use surveys typically do not provide time data for paid care services, so the estimates depend only on the time intensity of family care. With this caveat, the paper estimates that the care need in 2030 would require the equivalent of one-fifth to two-fifths of the paid labor force, assuming 40 weekly workhours. Using the projected 2030 mean wage for care and social service workers to estimate the hypothetical wage bill for these unpaid caregivers if they were paid, we obtain a value equivalent to 16 to 32 percent of GDP in the three countries. |
| Visualizing the invisible: class excursions to ignite children's enthusiasm for microbes | McGenity, Terry J.; Gessesse, Amare; Hallsworth, John E.; Garcia Cela, Esther; Verheecke-Vaessen, Carol; Wang, Fengping; Chavarría, Max; Haggblom, Max M.; Molin, Søren; Danchin, Antoine; Smid, Eddy J.; Lood, Cédric; Cockell, Charles S.; Whitby, | 2020-05-14 | Microb Biotechnol | https://www.ncbi.nlm.nih.gov/pubmed/32406115/; https://doi.org/10.1111/1751-7915.13576 | We have recently argued that, because microbes have pervasive – often vital – influences on our lives, and that therefore their roles must be taken into account in many of the decisions we face, society must become microbiology-literate, through the introduction of relevant microbiology topics in school curricula (Timmis et al. 2019. Environ Microbiol 21: 1513-1528). The current coronavirus pandemic is a stark example of why microbiology literacy is such a crucial enabler of informed policy decisions, particularly those involving preparedness of public-health systems for disease outbreaks and pandemics. However, a significant barrier to attaining widespread appreciation of microbial contributions to our well-being and that of the planet is the fact that microbes are seldom visible: most people are only peripherally aware of them, except when they fall ill with an infection. And it is disease, rather than all of the positive |

| Title | Authors | Date | Journal | URL | Abstract |
|---|---|---|---|---|---|
| | Corinne; Liu, Shuang-Jiang; Keller, Nancy P.; Stein, Lisa Y.; Bordenstein, Seth R.; Lal, Rup; Nunes, Olga C.; Gram, Lone; Singh, Brajesh K.; Webster, Nicole S.; Morris, Cindy; Sivinski, Sharon; Bindschedler, Saskia; Junier, Pilar; Antunes, André; Baxter, Bonnie K.; Scavone, Paola; Timmis, Kenneth | | | | activities mediated by microbes, that colours public perception of 'germs' and endows them with their poor image. It is imperative to render microbes visible, to give them life and form for children (and adults), and to counter prevalent misconceptions, through exposure to imagination-capturing images of microbes and examples of their beneficial outputs, accompanied by a balanced narrative. This will engender automatic mental associations between everyday information inputs, as well as visual, olfactory and tactile experiences, on the one hand, and the responsible microbes/microbial communities, on the other hand. Such associations, in turn, will promote awareness of microbes and of the many positive and vital consequences of their actions, and facilitate and encourage incorporation of such consequences into relevant decision-making processes. While teaching microbiology topics in primary and secondary school is key to this objective, a strategic programme to expose children directly and personally to natural and managed microbial processes, and the results of their actions, through carefully planned class excursions to local venues, can be instrumental in bringing microbes to life for children and, collaterally, their families. In order to encourage the embedding of microbiology-centric class excursions in current curricula, we suggest and illustrate here some possibilities relating to the topics of food (a favourite pre-occupation of most children), agriculture (together with horticulture and aquaculture), health and medicine, the environment and biotechnology. And, although not all of the microbially relevant infrastructure will be within reach of schools, there is usually access to a market, local food store, wastewater treatment plant, farm, surface water body, etc., all of which can provide opportunities to explore microbiology in action. If children sometimes consider the present to be mundane, even boring, they are usually excited with both the past and the future so, where possible, visits to local museums (the past) and research institutions advancing knowledge frontiers (the future) are strongly recommended, as is a tapping into the natural enthusiasm of local researchers to leverage the educational value of excursions and virtual excursions. Children are also fascinated by the unknown, so, paradoxically, the invisibility of microbes |

| Title | Authors | Date | Journal | URL | Abstract |
| --- | --- | --- | --- | --- | --- |
| | | | | | makes them especially fascinating objects for visualization and exploration. In outlining some of the options for microbiology excursions, providing suggestions for discussion topics and considering their educational value, we strive to extend the vistas of current class excursions and to: (i) inspire teachers and school managers to incorporate more microbiology excursions into curricula; (ii) encourage microbiologists to support school excursions and generally get involved in bringing microbes to life for children; (iii) urge leaders of organizations (biopharma, food industries, universities, etc.) to give school outreach activities a more prominent place in their mission portfolios, and (iv) convey to policymakers the benefits of providing schools with funds, materials and flexibility for educational endeavours beyond the classroom. |
| How to Best Protect People With Diabetes From the Impact of SARS-CoV-2: Report of the International COVID-19 and Diabetes Summit | Zhang, Jennifer Y.; Shang, Trisha; Ahn, David; Chen, Kong; Coté, Gerard; Espinoza, Juan; Mendez, Carlos E.; Spanakis, Elias K.; Thompson, Bithika; Wallia, Amisha; Wisk, Lauren E.; Kerr, David; Klonoff, David C. | 2021-01-21 | J Diabetes Sci Technol | https://www.ncbi.nlm.nih.gov/pubmed/33476193/; https://doi.org/10.1177/1932296820978399 | The coronavirus disease 2019 (COVID-19) pandemic caused by the severe acute respiratory syndrome coronavirus 2 (SARS-CoV-2) virus has rapidly involved the entire world and exposed the pressing need for collaboration between public health and other stakeholders from the clinical, scientific, regulatory, pharmaceutical, and medical device and technology communities. To discuss how to best protect people with diabetes from serious outcomes from COVID-19, Diabetes Technology Society, in collaboration with Sansum Diabetes Research Institute, hosted the "International COVID-19 and Diabetes Virtual Summit" on August 26-27, 2020. This unique, unprecedented real-time conference brought together physicians, scientists, government officials, regulatory experts, industry representatives, and people with diabetes from six continents to review and analyze relationships between COVID-19 and diabetes. Over 800 attendees logged in. The summit consisted of five sessions: (I) Keynotes, (II) Preparedness, (III) Response, (IV) Recovery, and (V) Surveillance; eight parts: (A) Background, (B) Resilience, (C) Outpatient Care, (D) Inpatient Care, (E) Resources, (F) High-Risk Groups, (G) Regulation, and (H) The Future; and 24 sections: (1) Historic Pandemics and Impact on Society, (2) Pathophysiology/Risk Factors for COVID-19, (3) Social Determinants of COVID-19, (4) Preparing for the Future, (5) Medications and Vaccines, (6) |

| Title | Authors | Date | Journal | URL | Abstract |
|---|---|---|---|---|---|
| | | | | | Psychology of Patients and Caregivers, (7) Outpatient Treatment of Diabetes Mellitus and Non-Pharmacologic Intervention, (8) Technology and Telehealth for Diabetes Outpatients, (9) Technology for Inpatients, (10) Management of Diabetes Inpatients with COVID-19, (11) Ethics, (12) Accuracy of Diagnostic Tests, (13) Children, (14) Pregnancy, (15) Economics of Care for COVID-19, (16) Role of Industry, (17) Protection of Healthcare Workers, (18) People with Diabetes, (19) International Responses to COVID-19, (20) Government Policy, (21) Regulation of Tests and Treatments, (22) Digital Health Technology, (23) Big Data Statistics, and 24) Patient Surveillance and Privacy. The two keynote speeches were entitled (1) COVID-19 and Diabetes—Meeting the Challenge and (2) Knowledge Gaps and Research Opportunities for Diabetes and COVID-19. While there was an emphasis on diabetes and its interactions with COVID-19, the panelists also discussed the COVID-19 pandemic in general. The meeting generated many novel ideas for collaboration between experts in medicine, science, government, and industry to develop new technologies and disease treatment paradigms to fight this global pandemic. |
| The Impact of COVID-19 on Health Behavior, Stress, Financial and Food Security among Middle to High Income Canadian Families with | Carroll, Nicholas; Sadowski, Adam; Laila, Amar; Hruska, Valerie; Nixon, Madeline; Ma, David W.L.; Haines, Jess | 2020-08-07 | Nutrients | https://www.ncbi.nlm.nih.gov/pubmed/32784530/; https://doi.org/10.3390/nu12082352 | The COVID-19 pandemic has disrupted many aspects of daily life. The purpose of this study was to identify how health behaviors, level of stress, financial and food security have been impacted by the pandemic among Canadian families with young children. Parents (mothers, n = 235 and fathers, n = 126) from 254 families participating in an ongoing study completed an online survey that included close and open-ended questions. Descriptive statistics were used to summarize the quantitative data and qualitative responses were analyzed using thematic analysis. More than half of our sample reported that their eating and meal routines have changed since COVID-19; most commonly reported changes were eating more snack foods and spending more time cooking. Screen time increased among 74% of mothers, 61% of fathers, and 87% of children and physical activity decreased among 59% of mothers, 52% of fathers, and 52% of children. Key factors influencing family stress include balancing work with childcare/homeschooling and financial instability. While some unhealthful |

| Title | Authors | Date | Journal | URL | Abstract |
|---|---|---|---|---|---|
| Young Children | | | | | behaviors appeared to have been exacerbated, other more healthful behaviors also emerged since COVID-19. Research is needed to determine the longer-term impact of the pandemic on behaviors and to identify effective strategies to support families in the post-COVID-19 context. |
| The 'Heart Kuznets Curve'? Understanding the relations between economic development and cardiac conditions | Nagano, Hitoshi; Puppim de Oliveira, Jose A.; Barros, Allan Kardec; Costa Junior, Altair da Silva | 2020-08-31 | World Development | https://www.ncbi.nlm.nih.gov/pubmed/32362711/; https://doi.org/10.1016/j.worlddev.2020.104953; https://api.elsevier.com/content/article/pii/S0305750X20300796; https://www.sciencedirect.com/science/article/pii/S0305750X20300796 | Abstract As countries turn wealthier, some health indicators, such as child mortality, seem to have well-defined trends. However, others, including cardiovascular conditions, do not follow clear linear patterns of change with economic development. Abnormal blood pressure is a serious health risk factor with consequences for population growth and longevity as well as public and private expenditure in health care and labor productivity. This also increases the risk of the population in certain pandemics, such as COVID-19. To determine the correlation of income and blood pressure, we analyzed time-series for the mean systolic blood pressure (SBP) of men's population (mmHg) and nominal Gross Domestic Product per capita (GDPPC) for 136 countries from 1980 to 2008 using regression and statistical analysis by Pearson's correlation (r). Our study finds a trend similar to an inverted-U shaped curve, or a 'Heart Kuznets Curve'. There is a positive correlation (increase GDPPC, increase SBP) in low-income countries, and a negative correlation in high-income countries (increase GDPPC, decrease SBP). As country income rises people tend to change their diets and habits and have better access to health services and education, which affects blood pressure. However, the latter two may not offset the rise in blood pressure until countries reach a certain income. Investing early in health education and preventive health care could avoid the sharp increase in blood pressure as countries develop, and therefore, avoiding the 'Heart Kuznets Curve' and its economic and human impacts. |
| Multisystem Resilience for Children and Youth in Disaster: | Masten, Ann S.; Motti-Stefanidi, Frosso | 2020-06-25 | Advers Resil Sci | https://www.ncbi.nlm.nih.gov/pubmed/32838305/; https://doi.org/10.100 | In the context of rising disasters worldwide and the challenges of the COVID-19 pandemic, this commentary considers the implications of findings in resilience science on children and youth for disaster preparation and response. The multisystem challenges posed by disasters are illustrated by the COVID-19 pandemic. We discuss the significance of disasters in the history of resilience |

| Title | Authors | Date | Journal | URL | Abstract |
|---|---|---|---|---|---|
| Reflections in the Context of COVID-19 | | | | 7/s42844-020-00010-w | science and the emergence of a unifying systems definition of resilience. Principles of a multisystem perspective on resilience and major findings on what matters for young people in disasters are delineated with reference to the pandemic. Striking parallels are noted in the psychosocial resilience factors identified at the level of individual children, families, schools, and communities. These parallels suggest that adaptive capacities associated with resilience in these interacting systems reflect interconnected networks and processes that co-evolved and may operate in concert. As resilience science moves toward integrated theory, knowledge, and applications in practice, particularly in disaster risk reduction and resilience promotion, more focus will be needed on multisystem and multidisciplinary research, communication, training, and planning. |
| Exploring resource scarcity and contextual influences on wellbeing among young refugees in Bidi Bidi refugee settlement, Uganda: findings from a qualitative study | Logie, Carmen H.; Okumu, Moses; Latif, Maya; Musoke, Daniel Kibuuka; Odong Lukone, Simon; Mwima, Simon; Kyambadde, Peter | 2021-01-07 | Confl Health | https://doi.org/10.1186/s13031-020-00336-3; https://www.ncbi.nlm.nih.gov/pubmed/33413546/ | BACKGROUND: Contextual factors including poverty and inequitable gender norms harm refugee adolescent and youths' wellbeing. Our study focused on Bidi Bidi refugee settlement that hosts more than 230,000 of Uganda's 1.4 million refugees. We explored contextual factors associated with wellbeing among refugee adolescents and youth aged 16–24 in Bidi Bidi refugee settlement. METHODS: We conducted 6 focus groups (n = 3: women, n = 3: men) and 10 individual interviews with young refugees aged 16–24 living in Bidi Bidi. We used physical distancing practices in a private outdoor space. Focus groups and individual interviews explored socio-environmental factors associated with refugee youth wellbeing. Focus groups were digitally recorded, transcribed verbatim, and coded by two investigators using thematic analysis. Analysis was informed by a social contextual theoretical approach that considers the interplay between material (resource access), symbolic (cultural norms and values), and relational (social relationships) contextual factors that can enable or constrain health promotion. RESULTS: Participants included 58 youth (29 men; 29 women), mean age was 20.9 (range 16–24). Most participants (82.8%, n = 48) were from South Sudan and the remaining from the Democratic Republic of Congo (17.2% [n = 10]). Participant narratives revealed the complex |

| Title | Authors | Date | Journal | URL | Abstract |
|---|---|---|---|---|---|
| | | | | | interrelationships between material, symbolic and relational contexts that shaped wellbeing. Resource constraints of poverty, food insecurity, and unemployment (material contexts) produced stress and increased sexual and gender-based violence (SGBV) targeting adolescent girls and women. These economic insecurities exacerbated inequitable gender norms (symbolic contexts) to increase early marriage and transactional sex (relational context) among adolescent girls and young women. Gendered tasks such as collecting water and firewood also increased SGBV exposure among girls and young women, and this was exacerbated by deforestation. Participants reported negative community impacts (relational context) of COVID-19 that were associated with fear and panic, alongside increased social isolation due to business, school and church closures. CONCLUSIONS: Resource scarcity produced pervasive stressors among refugee adolescents and youth. Findings signal the importance of gender transformative approaches to SGBV prevention that integrate attention to resource scarcity. These may be particularly relevant in the COVID-19 pandemic. Findings signal the importance of developing health enabling social contexts with and for refugee adolescents and youth. |
| Hand-Washing Practices among Adolescents Aged 12–15 Years from 80 Countries | Smith, Lee; Butler, Laurie; Tully, Mark A; Jacob, Louis; Barnett, Yvonne; López-Sánchez, Guillermo F.; López-Bueno, Rubén; Shin, Jae Il; McDermott, Daragh; Pfeifer, Briona A.; Pizzol, Damiano; Koyanagi, Ai | 2020-12-27 | Int J Environ Res Public Health | https://doi.org/10.3390/ijerph18010138; https://www.ncbi.nlm.nih.gov/pubmed/33375506/ | The objectives were to (1) assess the prevalence of hand-washing practices across 80 countries and (2) assess frequency of hand-washing practice by economic status (country income and severe food insecurity), in a global representative sample of adolescents. Cross-sectional data from the Global School-based Student Health Survey 2003–2017 were analyzed. Data on age, sex, hand-washing practices in the past 30 days, and severe food insecurity (i.e., proxy of socioeconomic status) were self-reported. Multivariable logistic regression and meta-analysis with random effects based on country-wise estimates were conducted to assess associations. Adolescents (n = 209,584) aged 12–15 years [mean (SD) age 13.8 (1.0) years; 50.9% boys] were included in the analysis. Overall, the prevalence of hand-washing practices were as follows: never/rarely washing hands before eating (6.4%), after using toilet (5.6%), or with soap (8.8%). The prevalence of never/rarely washing hands after using the |

| Title | Authors | Date | Journal | URL | Abstract |
| --- | --- | --- | --- | --- | --- |
| | | | | | toilet (10.8%) or with soap (14.3%) was particularly high in low-income countries. Severe food insecurity was associated with 1.34 (95%CI = 1.25–1.43), 1.61 (95%CI = 1.50–1.73), and 1.44 (95%CI = 1.35–1.53) times higher odds for never/rarely washing hands before eating, after using the toilet, and with soap, respectively. A high prevalence of inadequate hand washing practices was reported, particularly in low-income countries and those with severe food insecurity. In light of the present COVID-19 pandemic and the rapid expansion being observed in low- and middle-income locations, interventions that disseminate good hand-washing practices are urgently required. Such interventions may also have cross-over benefits in relation to other poor sanitation-related diseases. |
| Viral time capsule: a global photo-elicitation study of child and adolescent mental health professionals during COVID-19 | Herrington, Olivia D.; Clayton, Ashley; Benoit, Laelia; Prins-Aardema, Cecil; DiGiovanni, Madeline; Weller, Indigo; Martin, Andrés | 2021-02-02 | Child Adolesc Psychiatry Ment Health | https://doi.org/10.1186/s13034-021-00359-5; https://www.ncbi.nlm.nih.gov/pubmed/33531051/ | OBJECTIVE: To examine, through photo-elicitation, the personal and professional impact of the COVID-19 pandemic on mental health professionals working with children and adolescents around the globe. METHODS: We invited the submission of images collected about the pandemic between May and August 2020. We encouraged participants to yoke personal reflections or voice memos to their images. Using snowball sampling, we began with two invitations, including one to the graduates of a mentorship program continuously hosted since 2004 by the International Association of Child and Adolescent Psychiatry and Allied Professions (IACAPAP). We analyzed de-identified images and anonymized transcripts through iterative coding using thematic analysis informed by rich picture analysis and aided by NVivo software. RESULTS: We collected submissions from child and adolescent mental health professionals (n = 134) working in 54 countries spread across the five continents. We identified four overarching domains with component themes that revealed both the commonality and the uniqueness of the pandemic experience around the globe: (1) Place (adjusting to emptiness and stillness; shifting timeframes; blending of spaces); (2) Person (disruption to life rhythms; emotional toll; positives of the pandemic); (3) Profession (changing practices; outreach efforts; guild pride—and guilt); and (4) Purpose (from pandemic to syndemic; from lamenting to |

| Title | Authors | Date | Journal | URL | Abstract |
|---|---|---|---|---|---|
| | | | | | embracing; planning toward a better tomorrow). CONCLUSIONS: Photo-elicitation provided a disarming and efficient means to learn about individual, regional, and global similarities and differences regarding the professionals charged with addressing the mental health needs of children and adolescents around the globe. These findings may help inform practice changes in post-pandemic times. |
| Covid-19 deaths in Africa: prospective systematic postmortem surveillance study | Mwananyanda, Lawrence; Gill, Christopher J; MacLeod, William; Kwenda, Geoffrey; Pieciak, Rachel; Mupila, Zachariah; Lapidot, Rotem; Mupeta, Francis; Forman, Leah; Ziko, Luunga; Etter, Lauren; Thea, Donald | 2021-02-17 | BMJ | https://www.ncbi.nlm.nih.gov/pubmed/33597166/; https://doi.org/10.1136/bmj.n334 | OBJECTIVE: To directly measure the fatal impact of coronavirus disease 2019 (covid-19) in an urban African population. DESIGN: Prospective systematic postmortem surveillance study. SETTING: Zambia's largest tertiary care referral hospital. PARTICIPANTS: Deceased people of all ages at the University Teaching Hospital morgue in Lusaka, Zambia, enrolled within 48 hours of death. MAIN OUTCOME MEASURE: Postmortem nasopharyngeal swabs were tested via reverse transcriptase quantitative polymerase chain reaction (PCR) against severe acute respiratory syndrome coronavirus 2 (SARS-CoV-2). Deaths were stratified by covis-19 status, location, age, sex, and underlying risk factors. RESULTS: 372 participants were enrolled between June and September 2020; PCR results were available for 364 (97.8%). SARS-CoV-2 was detected in 58/364 (15.9%) according to the recommended cycle threshold value of <40 and in 70/364 (19.2%) when expanded to any level of PCR detection. The median age at death among people with a positive test for SARS-CoV-2 was 48 (interquartile range 36-72) years, and 69% (n=48) were male. Most deaths in people with covid-19 (51/70; 73%) occurred in the community; none had been tested for SARS-CoV-2 before death. Among the 19/70 people who died in hospital, six were tested before death. Among the 52/70 people with data on symptoms, 44/52 had typical symptoms of covid-19 (cough, fever, shortness of breath), of whom only five were tested before death. Covid-19 was identified in seven children, only one of whom had been tested before death. The proportion of deaths with covid-19 increased with age, but 76% (n=53) of people who died were aged under 60 years. The five most common comorbidities among people who died with covid-19 were tuberculosis (22; 31%), hypertension (19; 27%), |

| Title | Authors | Date | Journal | URL | Abstract |
|---|---|---|---|---|---|
| | | | | | HIV/AIDS (16; 23%), alcohol misuse (12; 17%), and diabetes (9; 13%). CONCLUSIONS: Contrary to expectations, deaths with covid-19 were common in Lusaka. Most occurred in the community, where testing capacity is lacking. However, few people who died at facilities were tested, despite presenting with typical symptoms of covid-19. Therefore, cases of covid-19 were under-reported because testing was rarely done not because covid-19 was rare. If these data are generalizable, the impact of covid-19 in Africa has been vastly underestimated. |
| Data-informed recommendations for services providers working with vulnerable children and families during the COVID-19 pandemic | Wilke, Nicole Gilbertson; Howard, Amanda Hiles; Pop, Delia | 2020-07-30 | Child Abuse Negl | https://www.sciencedirect.com/science/article/pii/S0145213420302970?v=s5; https://doi.org/10.1016/j.chiabu.2020.104642; https://api.elsevier.com/content/article/pii/S0145213420302970; https://www.ncbi.nlm.nih.gov/pubmed/32753231/ | BACKGROUND: The COVID-19 pandemic and associated response measures have led to unprecedented challenges for service providers working with vulnerable children and families around the world. OBJECTIVE: The goal of the present study was to better understand the impact of the pandemic and associated response measures on vulnerable children and families and provide data-informed recommendations for public and private service providers working with this population. PARTICIPANTS AND SETTING: Representatives from 87 non-government organizations (NGOs) providing a variety of direct services (i.e. residential care, family preservation, foster care, etc.) to 454,637 vulnerable children and families in 43 countries completed a brief online survey. METHODS: Using a mixed methods design, results examined 1) ways in which children and families have been directly impacted by COVID-19, 2) the impact of the pandemic on services provided by NGOs, 3) government responses and gaps in services for this population during the pandemic, and 4) strategies that have been effective in filling these gaps. RESULTS: Data revealed that the pandemic and restrictive measures were associated with increased risk factors for vulnerable children and families, including not having access to vital services. The NGOs experienced government restrictions, decreased financial support, and inability to adequately provide services. Increased communication and supportive activities had a positive impact on both NGO staff and the families they serve. CONCLUSIONS: Based on the findings, ten recommendations were made for service providers working with vulnerable children and families during the COVID-19 pandemic. |

| Title | Authors | Date | Journal | URL | Abstract |
|---|---|---|---|---|---|
| Baby pangolins on my plate: possible lessons to learn from the COVID-19 pandemic | Volpato, Gabriele; Fontefrancesco, Michele F.; Gruppuso, Paolo; Zocchi, Dauro M.; Pieroni, Andrea | 2020-04-21 | J Ethnobiol Ethnomed | https://doi.org/10.1186/s13002-020-00366-4; https://www.ncbi.nlm.nih.gov/pubmed/32316979/ | The Journal of Ethnobiology and Ethnomedicine (JEET), throughout its 15 years of existence, has tried to provide a respected outlet for scientific knowledge concerning the inextricable links between human societies and nature, food, and health. Ethnobiology and ethnomedicine-centred research has moved at the (partially artificial and fictitious) interface between nature and culture and has investigated human consumption of wild foods and wild animals, as well as the use of wild animals or their parts for medicinal and other purposes, along with the associated knowledge, skills, practices, and beliefs. Little attention has been paid, however, to the complex interplay of social and cultural reasons behind the increasing pressure on wildlife. The available literature suggest that there are two main drivers that enhance the necessary conditions for infectious diseases to cross the species barrier from wild animals to humans: (1) the encroachment of human activities (e.g., logging, mining, agricultural expansion) into wild areas and forests and consequent ecological disruptions; and, connected to the former, (2) the commodification of wild animals (and natural resources in general) and an expanding demand and market for wild meat and live wild animals, particularly in tropical and sub-tropical areas. In particular, a crucial role may have been played by the bushmeat-euphoria and attached elitist gastronomies and conspicuous consumption phenomena. The COVID-19 pandemic will likely require ethnobiologists to reschedule research agendas and to envision new epistemological trajectories aimed at more effectively mitigating the mismanagement of natural resources that ultimately threats our and other beings' existence. |
| Adapting HIV services for pregnant and breastfeeding women, infants, | Vrazo, Alexandra C; Golin, Rachel; Fernando, Nimasha B; Killam, Wm P; Sharifi, Sheena; Phelps, B Ryan; Gleason, Megan M; Wolf, Hilary T; Siberry, | 2020-09-01 | J Int AIDS Soc | https://doi.org/10.1002/jia2.25622; https://www.ncbi.nlm.nih.gov/pubmed/32996705/ | INTRODUCTION: The COVID-19 pandemic has impacted global health service delivery, including provision of HIV services. Countries with high HIV burden are balancing the need to minimize interactions with health facilities to reduce the risk of COVID-19 transmission, while delivering uninterrupted essential HIV prevention, testing and treatment services. Many of these adaptations in resource-constrained settings have not adequately accounted for the needs of pregnant and breastfeeding women, infants, children and adolescents. We propose whole- |

| Title | Authors | Date | Journal | URL | Abstract |
|---|---|---|---|---|---|
| children, adolescents and families in resource-constrained settings during the COVID-19 pandemic | George K; Srivastava, Meena | | | | family, tailored programme adaptations along the HIV clinical continuum to protect the programmatic gains made in services. DISCUSSION: Essential HIV case-finding services for pregnant and breastfeeding women and children should be maintained and include maternal testing, diagnostic testing for infants exposed to HIV, index testing for children whose biological parents or siblings are living with HIV, as well as for children/adolescents presenting with symptoms concerning for HIV and comorbidities. HIV self-testing for children two years of age and older should be supported with caregiver and provider education. Adaptations include bundling services in the same visit and providing testing outside of facilities to the extent possible to reduce exposure risk to COVID-19. Virtual platforms can be used to identify vulnerable children at risk of HIV infection, abuse, harm or violence, and link them to necessary clinical and psychosocial support services. HIV treatment service adaptations for families should focus on family based differentiated service delivery models, including community-based ART initiation and multi-month ART dispensing. Viral load monitoring should not be a barrier to transitioning children and adolescents experiencing treatment failure to more effective ART regimens, and viral load monitoring for pregnant and breastfeeding women and children should be prioritized and bundled with other essential services. CONCLUSIONS: Protecting pregnant and breastfeeding women, infants, children and adolescents from acquiring SARS-CoV-2 while sustaining essential HIV services is an immense global health challenge. Tailored, family friendly programme adaptations for case-finding, ART delivery and viral load monitoring for these populations have the potential to limit SARS-CoV-2 transmission while ensuring the continuity of life-saving HIV case identification and treatment efforts. |
| Assessing Food Poverty, Vulnerability and Food | Bidisha, Sayema Haque; Mahmood, Tanveer; Hossain, Md. Biplob | 2021-01-04 | Soc Indic Res | https://www.ncbi.nlm.nih.gov/pubmed/33424082/; https://doi.org/10.100 | There is no denying the fact that, for a developing country like Bangladesh, the economic consequences of lockdown for containing COVID-19 pandemic can be far reaching affecting livelihoods of millions of households. Given that the share of food consumption expenditure to total expenditure is higher in the lower income groups of Bangladesh, this shock is expected to directly affect |

| Title | Authors | Date | Journal | URL | Abstract |
|---|---|---|---|---|---|
| Consumption Inequality in the Context of COVID-19: A Case of Bangladesh | | | | 7/s11205-020-02596-1 | affordability of consumption of basic food items of these households. Using nationally representative household survey data of Bangladesh, and while following the Feasible Generalized Least Square method, this paper attempts to examine food poverty, food consumption inequality along with vulnerability to food poverty of households and explores the importance of different socio-demographic and environmental factors in this connection. Our estimation reflects that, greater percentage of households with young children or with elderly people are found to suffer high food vulnerability. In addition, households in environmentally endangered regions e.g. drought prone areas or river erosion affected places are more food vulnerable than those in other parts of the country. Certain occupation groups e.g. day labourer and self-employed are found to be highly vulnerable to food poverty while according to our decomposition analysis of food consumption inequality, area of residence (urban vs. rural) is expected to cause sizable inequality in food consumption. This study can therefore, help in identifying food vulnerable households for government's social protection programs and COVID-19 incentive packages, and thereby can contribute towards designing effective poverty reduction strategies. |
| Epidemiologic, clinical, and laboratory findings of the COVID-19 in the current pandemic: systematic review and meta-analysis | Xie, Yewei; Wang, Zaisheng; Liao, Huipeng; Marley, Gifty; Wu, Dan; Tang, Weiming | 2020-08-31 | BMC Infect Dis | https://doi.org/10.1186/s12879-020-05371-2; https://www.ncbi.nlm.nih.gov/pubmed/32867706/ | BACKGROUND: The COVID-19 pandemic has affected the world deeply, with more than 14,000,000 people infected and nearly 600,000 deaths. This review aimed to summarize the epidemiologic traits, clinical spectrum, CT results and laboratory findings of the COVID-19 pandemic. METHODS: We scoped for relevant literatures published during 1st December 2019 to 16th July 2020 based on three databases using English and Chinese languages. We reviewed and analyzed the relevant outcomes. RESULTS: The COVID-19 pandemic was found to have a higher transmission rate compared to SARS and MERS and involved 4 stages of evolution. The basic reproduction number (R(0)) is 3.32 (95% CI:3.24–3.39), the incubation period was 5.24 days (95% CI:3.97–6.50, 5 studies) on average, and the average time for symptoms onset varied by countries. Common clinical spectrums identified included fever (38.1–39.0 °C), cough and fatigue, with Acute Respiratory Distress Syndrome (ARDS) being the |

| Title | Authors | Date | Journal | URL | Abstract |
|---|---|---|---|---|---|
| | | | | | most common complication reported. Body temperatures above 39.0 °C, dyspnea, and anorexia were more common symptoms in severe patients. Aged over 65 years old, having co-morbidities, and developing complications were the commonest high-risk factors associated with severe conditions. Leucopenia and lymphopenia were the most common signs of infection while liver and kidney damage were rare but may cause bad outcomes for patients. The bilateral, multifocal Ground-Glass Opacification (GGO) on peripheral, and the consolidative pulmonary opacity were the most frequent CT results and the tendency of mortality rates differed by region. CONCLUSIONS: We provided a bird's-eye view of the COVID-19 during the current pandemic, which will help better understanding the key traits of the disease. The findings could be used for disease's future research, control and prevention. |
| Kidney disease and electrolytes in COVID-19: more than meets the eye | Carriazo, Sol; Kanbay, Mehmet; Ortiz, Alberto | 2020-07-16 | Clin Kidney J | https://doi.org/10.1093/ckj/sfaa112; https://www.ncbi.nlm.nih.gov/pubmed/32699613/ | COVID-19 is a global pandemic fuelled in some countries by government actions. The current issue of Clinical Kidney Journal presents 15 articles on COVID-19 and kidney disease from three continents, providing a global perspective of the impact of severe acute respiratory syndrome coronavirus 2 on electrolytes and different kidney compartments (glomeruli, tubules and vascular compartments) and presenting clinically as a syndrome of inappropriate antidiuretic hormone secretion, acute kidney injury, acute kidney disease, collapsing glomerulopathy and thrombotic microangiopathy, among others, in the context of a brand-new cardiorenal syndrome. Kidney injury may need acute dialysis that may overwhelm haemodialysis (HD) and haemofiltration capabilities. In this regard, acute peritoneal dialysis (PD) may be lifesaving. Additionally, pre-existent chronic kidney disease increases the risk of more severe COVID-19 complications. The impact of COVID-19 on PD and HD patients is also discussed, with emphasis on preventive measures. Finally, current therapeutic approaches and potential future therapeutic approaches undergoing clinical trials, such as complement targeting by eculizumab, are also presented. |

| Title | Authors | Date | Journal | URL | Abstract |
|---|---|---|---|---|---|
| Covid-19, ACE2 and the kidney | Hardenberg, Jan-Hendrik; Luft, Friedrich C. | 2020-08-02 | Acta Physiol (Oxf) | https://www.ncbi.nlm.nih.gov/pubmed/32662161/; https://doi.org/10.1111/apha.13539 | We are confronted with the most dramatic pandemic world-wide for the past 100 years. We are armed "to-the-teeth" compared to 1918, we know the agent, the genomic sequence, the bodily entry, the proliferation rate, the damage pathogenesis, and the very nature of our enemy. We can identify its bodily presence and our resistance to it in terms of neutralizing antibody production. Nonetheless, the disease has laid lame the great nations of the current world and crippled the less fortunate countries. The primary disease features are not the kidney. However, the entry point has much to do with renal and cardiovascular disease. The kidney is a common target of corona-virus (SARS-CoV2) disease; the longer-term consequences could be as well. |
| Overcrowding and Exposure to Secondhand Smoke Increase Risk for COVID-19 Infection Among Latinx Families in Greater San Francisco Bay Area | DeCastro Mendez, A.; Escobar, M.; Romero Encinas, M.; Wojcicki, J. | 2021-01-20 | | https://doi.org/10.1101/2021.01.19.21250139; http://medrxiv.org/cgi/content/short/2021.01.19.21250139v1?rss=1 | Background: The novel coronavirus (COVID-19) has disproportionately impacted the Latinx community in the United States. Environmental risk factors, including community level pollution burden and exposure to smoking and secondhand smoke, have not been evaluated in relation to risk for infection with COVID-19. Methods: We evaluated self-reported infection rates of COVID-19 in three, preexisting, longitudinal, Latinx family cohorts in the San Francisco Bay Area from May through September 2020 (N=383 households, 1,875 people). All households were enrolled during pregnancy and postpartum at Zuckerberg San Francisco General Hospital (ZSFG) and UCSF Benioff before the pandemic. For the COVID-19 sub-study, participants responded to a 15-minute telephonic interview where we assessed food consumption patterns, housing and employment status, and history of COVID-19 infection based on community and hospital-based testing. We also evaluated secondhand smoke exposure based on previously collected data. Environmental pollution exposure was determined from census tract residence using California EnviroScreen 2.0 data. Non-parametric tests were used to assess possible associations and multiple logistic regression analyses to determine independent predictors of COVID-19 infection. Results: In the combined Latinx, Eating and Diabetes Cohort (LEAD) and Hispanic, Eating and Nutrition (HEN) cohorts there was a 7.6% household infection rate (14/183) with a lower rate of 3.5% (7/200) in the Telomeres at Birth |

| Title | Authors | Date | Journal | URL | Abstract |
|---|---|---|---|---|---|
| | | | | | (TAB) cohort. Larger household size increased risk for infection (OR, 1.43 (95%CI 1.10-1.87)) in the combined LEAD/HEN cohorts and increasing number of children trended towards significance in the TAB cohort (OR 1.82, 95% CI 0.98-3.37). Any exposure to secondhand smoke in the household also trended towards increasing risk after adjusting for household size and other exposures (OR 3.20, 95%CI 0.80-12.73) and (OR 4.37, 95% CI 0.80-23.70). We did not find any associations between neighborhood pollution level and COVID-19 infection based on census track and risk of infection. Furthermore, we found weak evidence between dietary exposure and risk of COVID-19 infection after adjusting for possible confounders. Conclusion: Crowding as indicated by household size increases risk for COVID-19 infection in Latinx families. Exposure to any secondhand smoke may also increase risk for COVID-19 through increased coughing and risk for respiratory impairment. Public policy and health interventions need to ensure that multi-unit residential complexes prevent any exposure to secondhand smoke. |

#### Hand- and Snowball Search

| Title | Authors | Date | Journal | URL | Abstract |
|---|---|---|---|---|---|
| Child malnutrition and COVID-19: the time to act is now | Fore, H. H., Dongyu, Q., Beasley, D. M., & Ghebreyesus, T. A. | 2020-08-22 | The Lancet | https://www.thelancet.com/article/S0140-6736(20)31648-2/fulltext | The COVID-19 pandemic is undermining nutrition across the world, particularly in low-income and middle-income countries (LMICs).1 The worst consequences are borne by young children. Some of the strategies to respond to COVID-19—including physical distancing, school closures, trade restrictions, and country lockdowns—are impacting food systems by disrupting the production, transportation, and sale of nutritious, fresh, and affordable foods, forcing millions of families to rely on nutrient-poor alternatives. Strained health systems and interruptions in humanitarian response are eroding access to essential and often life-saving nutrition services.2 Social |

| | | | | | |
|---|---|---|---|---|---|
| | | | | | protection systems in many LMICs are overloaded as vulnerable families struggle to access the food and services they need in the context of an economic downturn. |
| Impacts of COVID-19 on childhood malnutrition and nutrition-related mortality | Headey, D., Heidkamp, R., Osendarp, S., Ruel, M., Scott, N., Black, R., ... & Walker, N. | 2020-07-27 | The Lancet | https://doi.org/10.1016/S0140-6736(20)31647-0 | The unprecedented global social and economic crisis triggered by the COVID-19 pandemic poses grave risks to the nutritional status and survival of young children in low-income and middle-income countries (LMICs). Of particular concern is an expected increase in child malnutrition, including wasting, due to steep declines in household incomes, changes in the availability and affordability of nutritious foods, and interruptions to health, nutrition, and social protection services.1 |
| Is the effect of COVID-19 on children underestimated in low-and middle-income countries? | Simba, J., Sinha, I., Mburugu, P., Agweyu, A., Emadau, C., Akech, S., ... & English, M. | 2020-06-18 | Acta Paediatrica | https://doi.org/10.1111/apa.15419 | The COVID-19 pandemic has had a huge impact on health and society, worldwide. While most high-income countries are felt to be reaching their COVID-19 peak, most low- and middle-income countries (LMICs), particularly sub-Saharan countries, are anticipating an exponential growth of cases.1 Overall it has been documented that children are less affected.2 However, in this commentary we describe how in Kenya, a LMIC in sub-Saharan Africa, COVID-19 is likely to have far-reaching direct and indirect implications on children. Of all confirmed cases of COVID-19, only around 1%-5% are children.2 In Kenya, the first case of COVID-19 was diagnosed on 13th March 2020; one month later, six children aged between one and 15 years (2.7% out of 216 total cases) have been diagnosed with the infection according to the Ministry of Health Kenya, COVID-19 Daily Situation Report as at 13th April 2020. Although most have had mild illness, there has been one reported death of a child with underlying comorbidity. As testing for COVID-19 in children is very limited, and children being excluded from mass testing, it is difficult to be confident in the numbers. Collecting nasopharyngeal or oropharyngeal swabs requires the patient to be cooperative, which is difficult to achieve in young children, as well as acute respiratory infections (ARIs), are so common in children that index of suspicion of COVID-19 is low, these pose unique challenges in children. It is therefore imperative that other methods of estimating the burden of COVID-19 in LMICs may have to be utilised including syndromic surveillance as well as modelling data. However, modelling in COVID-19 has led to different predictions depending on assumptions made.3 It is therefore difficult to tell which path the pandemic will follow in reality. |

| | | | | | |
|---|---|---|---|---|---|
| Things must not fall apart: the ripple effects of the COVID-19 pandemic on children in sub-Saharan Africa | Modupe Coker, Morenike O. Folayan, Ian C. Michelow, Regina E. Oladokun, Nguavese Torbunde & Nadia A. Sam-Agudu | 2020-09-24 | Nature - Pediatric Research | https://www.nature.com/articles/s41390-020-01174-y | Zero to 19 year-old children in sub-Saharan Africa bear a disproportionate proportion of the global burden of communicable and non-communicable diseases. Significant public health gains have been made in the fight against these diseases, however, factors such as underequipped health systems, disease outbreaks, conflict, and political instability continue to challenge prevention and control. The novel coronavirus disease (COVID-19) pandemic caused by severe acute respiratory syndrome coronavirus 2 (SARS-CoV-2) introduces new challenges to public health programs in sub-Saharan Africa. Of particular concern are programs targeting major conditions among children, such as undernutrition, vaccine-preventable pneumonia and diarrhea, malaria, tuberculosis, HIV, and sickle cell disease. This article focuses on the impact of the COVID-19 pandemic on child health in sub-Saharan Africa. We review the epidemiology of major pediatric diseases and, referencing modeling projections, discuss the short- and long-term impact of the pandemic on major disease control. We deliberate on potential complications of SARS-CoV-2 co-infections/co-morbidities and identify critical social and ethical issues. Furthermore, we highlight the paucity of COVID-19 data and clinical trials in this region and the lack of child participants in ongoing studies. Lastly, approaches and interventions to mitigate the pandemic's impact on child health outcomes are discussed. |
| Challenges of COVID-19 in children in low- and middle-income countries | Zar HJ, Dawa J, Fischer GB, Castro-Rodriguez JA. | 2020-06-25 | Paediatr Respir Rev. | https://doi.org/10.1016/j.prrv.2020.06.016 | As the coronavirus pandemic extends to low and middle income countries (LMICs), there are growing concerns about the risk of coronavirus disease (COVID-19) in populations with high prevalence of comorbidities, the impact on health and economies more broadly and the capacity of existing health systems to manage the additional burden of COVID-19. The direct effects of COVID are less of a concern in children, who seem to be largely asymptomatic or to develop mild illness as occurs in high income countries; however children in LMICs constitute a high proportion of the population and may have a high prevalence of risk factors for severe lower respiratory infection such as HIV or malnutrition. Further diversion of resources from child health to address the pandemic among adults may further impact on care for children. Poor living conditions in LMICs including lack of sanitation, running water and overcrowding may facilitate transmission of SARS-CoV-2. The indirect effects of the pandemic on child health are of considerable concern, including |

| | | | | | |
|---|---|---|---|---|---|
| | | | | | increasing poverty levels, disrupted schooling, lack of access to school feeding schemes, reduced access to health facilities and interruptions in vaccination and other child health programs. Further challenges in LMICs include the inability to implement effective public health measures such as social distancing, hand hygiene, timely identification of infected people with self-isolation and universal use of masks. Lack of adequate personal protective equipment, especially N95 masks is a key concern for health care worker protection. While continued schooling is crucial for children in LMICs, provision of safe environments is especially challenging in overcrowded resource constrained schools. The current crisis is a harsh reminder of the global inequity in health in LMICs. The pandemic highlights key challenges to the provision of health in LMICs, but also provides opportunities to strengthen child health broadly in such settings. |
| A Crisis within a Crisis: COVID-19 and Hunger in African Children | Aborode AT, Ogunsola SO, Adeyemo AO. A | 2020-01 | Am J Trop Med Hyg. | https://doi.org/10.4269/ajtmh.20-1213 | The WHO recently expressed concern at the potential impact of COVID-19 on hunger, which is likely to exacerbate the already considerable burden of malnutrition of Africa. The impact of the disease is expected to be greater among those grappling with malnutrition, whereas widespread hunger and malnutrition will likely increase because of movement restrictions. COVID-19 is unfolding in Africa against a backdrop of worrying levels of hunger and undernourishment which could worsen as the virus threatens livelihoods and household economies. The perspective piece addresses the crisis within crisis of COVID-19 and hunger on the well-being of children in Africa. |
| The risk to child nutrition during and after COVID-19 pandemic: what to expect and how to respond | Ntambara J, Chu M. | 2021-04-13 | Public Health Nutr. | https://doi.org/10.1017/s1368980021001610 | Objective: This study aimed to address the key areas of concern for child nutrition, both during and after the COVID-19 pandemic, and proposes strategic responses to reduce child undernutrition in the short and long term.<br>Design: A descriptive literature review was performed. The search of the literature was conducted through using electronic databases including PubMed, Web of science, google scholar, and Cochrane library.<br>Setting: A wide range of published articles focused on child malnutrition were reviewed. |

| | | | | | |
|---|---|---|---|---|---|
| | | | | | Participants: The study was focused on children especially those under five years.<br>Results: This study proposes strategic responses to reduce child undernutrition. These responses include strengthening access to community-based nutrition services that support the early detection and treatment of undernourished children and emergency food distribution, including fortified foods with vitamins and minerals, to vulnerable households, particularly those with children under five years. Moreover, counseling and promotion programs should be reinforced to revitalize community nutrition education in areas such as gestation, exclusive breastfeeding, and complementary feeding, and hygienic practices involving handwashing, proper sanitation, and other basic behavioral changes.<br>Conclusions: The COVID-19 pandemic has affected many countries especially those in the regions of South Asia and sub-Saharan Africa in which there has been an ongoing burden of child undernutrition. However, malnutrition is preventable and can be eliminated through a multisectoral strategic approach. The effective execution of a multisectoral approach toward preventing childhood malnutrition will require not only a financial investment but also the collective efforts from different ministries of the governments, UN-affiliated agencies, and nongovernmental organizations. |
| Child Undernutrition in Sudan: The Social and Economic Impact and Future Perspectives | Abu-Fatima O, Abbas AAG, Racalbuto V, Smith L, Pizzol D. | 2021-03-01 | Am J Trop Med Hyg. | https://doi.org/10.4269/ajtmh.20-1251 | The nutrition situation in Sudan is one of the worst in northeast Africa and it is characterized by persistently high levels of acute and chronic malnutrition that have increased over the last two decades. The underlying causes of malnutrition are multi-sectoral and are mainly due to inequalities, inadequate food practices, and limited access to healthcare services. Based on the report The Economic and Social Impacts of Child Undernutrition in Sudan, this study assesses the impact that malnutrition has on health, education, and productivity in Sudan. The country is estimated to have lost an equivalent of about 11.6 billion Sudanese pound in 2014, which represented 2.6% of the gross domestic product (GDP). Productivity-related losses contributed the largest costs at 1.5% of GDP followed by health and education sectors at 1.1% and 0.1%, respectively. In 2020, the outbreak of the COVID-19 pandemic further exposed the fragility of Sudan's health, social, and |

| | | | | | |
|---|---|---|---|---|---|
| | | | | | economic system. It is mandatory that all stakeholders address child nutrition as a main concern and stunting is incorporated in the center of the development agenda. In particular, the national development frameworks should be updated to ensure the reduction of the stunting prevalence and to put in place a comprehensive multi-sectoral nutrition policy, strategy, and plan of action. |
| The implications of COVID-19 for the children of Africa. | Mustafa, F., & J Green, R. | 2020-06 | Afr. med. j. | http://dx.doi.org/10.7196/SAMJ.2020.v110i6.14824 | COVID-19, and the novel coronavirus causing it, has been declared a pandemic by the World Health Organization. It presents with signs and symptoms of respiratory illness, and other nonspecific symptoms. These symptoms may be mild enough to go unnoticed, or severe enough to overwhelm a healthcare system in a First-World country. It is an emerging problem that can potentially put intolerable strain on a health system that is fragile and likely to collapse, such as those that exist in Africa. Extraordinary times like these require ingenious statesmanship and astutely calculated plans to see a nation emerge through the crisis. And in such a crisis, special attention needs to be directed to the healthcare system, where medical attention, equipment and interventions need to be carefully rationed.<br><br>To date, sub-Saharan Africa has not seen the devastation wrought on the Northern Hemisphere. That may be fortuitous, we may be lucky, it may just be coming.<br><br>In addition, SARS-CoV-2 has left children largely unaffected by disease.[1] They may be the silent carriers, which could have its own awful psychological impact. But whether or not COVID-19 ultimately affects the children of Africa directly, it will leave them scarred, worse off, and still facing a burden of infectious diseases not seen anywhere else in the world. |
| Assessing the Zero Hunger Target Readiness in Africa in the | Otekunrin, O. A., Otekunrin, O. A., Fasina, F. O., Omotayo, A. | 2020-07-15 | Caraka Tani: Journal of Sustainable | https://scholar.googleusercontent.com/scholar?q=cache:Dh_gtOWa9tUJ:scholar.googl | Sustainable Development Goal 2 (SDG 2) is hinged on achieving zero hunger target globally, by the year 2030. Many developing countries, especially African countries, are faced with extreme hunger often caused or compounded by bad governance, conflicts and climate change. In this paper, we assess Africa's readiness towards attaining the zero hunger target by 2030 in the face of COVID-19 pandemic. Patterns |

| | | | | | |
|---|---|---|---|---|---|
| face of COVID-19 Pandemic | O., & Akram, M. | | Agriculture | e.com/+covid-19+and+malnutrition+in+african+children&hl=de&as_sdt=0,5 | of Global Hunger Index (GHI) and each of its indicators across Africa are compared before the pandemic (2000-2019). The effect of the pandemic on the hunger situation in Africa is discussed highlighting the mitigating measures put in place by selected African governments. We found that most African countries have recorded steady reduction in their child mortality rates but high prevalence of undernourishment, stunting and child wasting indicate significant challenges hampering the achievement of the zero hunger target. The study recommends that African governments should prioritise sustainable agricultural practices while serious attention should be given to the formulation and implementation of policies that reduce hunger in the face the COVID-19 pandemic. |
| COVID-19 implications on household income and food security in Kenya and Uganda: Findings from a rapid assessment. | Kansiime, M. K., Tambo, J. A., Mugambi, I., Bundi, M., Kara, A., & Owuor, C. | 2021-01 | World development | https://doi.org/10.1016/j.worlddev.2020.105199 | This study assessed implications of the Coronavirus Disease 19 (COVID-19) pandemic on household income and food security in two East African countries – Kenya and Uganda, using online survey data from 442 respondents. Results show that more than two-thirds of the respondents experienced income shocks due to the COVID-19 crisis. Food security and dietary quality worsened, as measured by the food insecurity experience scale and the frequency of consumption of nutritionally-rich foods. The proportion of food insecure respondents increased by 38% and 44% in Kenya and Uganda respectively, and in both countries, the regular consumption of fruits decreased by about 30% during the COVID-19 pandemic, compared to a normal period (before the pandemic). Results from probit regressions show that the income-poor households and those dependent on labour income were more vulnerable to income shock, and had poorer food consumption during the COVID-19 pandemic compared to other respondent categories. As such, they were more likely to employ food-based coping strategies compared to those pursuing alternative livelihoods, who generally relied on savings. Farmers were less likely to experience worsened food security compared to other respondent categories who depended to a great extent on market sources for food. In both countries, participation in national social security schemes was less likely to mitigate respondents' income shock during the COVID-19 period. Conversely, membership in savings and loan groups was correlated with less likelihood of suffering income |

| | | | | | |
|---|---|---|---|---|---|
| | | | | | shocks and reduction in food consumption. The results suggest that ongoing and future government responses should focus on structural changes in social security by developing responsive packages to cushion members pushed into poverty by such pandemics while building strong financial institutions to support the recovery of businesses in the medium term, and ensuring the resilience of food supply chains particularly those making available nutrient-dense foods. |
| Food security and welfare changes under COVID-19 in Sub-Saharan Africa: Impacts and responses in Kenya. | Nechifor, V., Ramos, M. P., Ferrari, E., Laichena, J., Kihiu, E., Omanyo, D., ... & Kiriga, B. | 2021-03 | World development | https://doi.org/10.1016/j.gfs.2021.100514 | The COVID-19 pandemic has affected all Sub-Saharan economies through a multitude of impact channels. The study determines the medium-term macroeconomic outcomes of the pandemic on the Kenyan economy and links the results with a detailed food security and nutrition microsimulation module. It thus evaluates the effectiveness of the adopted government measures to reduce the negative outcomes on food security and to enable economic recovery at aggregate, sectoral and household levels. Through income support measures, the food sector and food demand partially recover. However, 1.3% of households still fall below calorie intake thresholds, many of which are in rural areas. Results also indicate that the state of food security in Kenya remains vulnerable to the evolution of the pandemic abroad. |
