## Supplemental File 3 - Literature Snowball and Hand Search for "The COVID-19 pandemic and child malnutrition in sub-Saharan Africa: A scoping review"

| | | | | | |
|---|---|---|---|---|---|
| Covid-19 lockdowns, income distribution, and food security: An analysis for South Africa | Arndt, C., Davies, R., Gabriel, S., Harris, L., Makrelov, K., Robinson, S., ... & Anderson, L. | 2020-09 | Global Food Security | https://doi.org/10.1016/j.gfs.2020.100410 | Absent vaccines and pharmaceutical interventions, the only tool available to mitigate its demographic effects is some measure of physical distancing, to reduce contagion by breaking social and economic contacts. Policy makers must balance the positive health effects of strong distancing measures, such as lockdowns, against their economic costs, especially the burdens imposed on low income and food insecure households. The distancing measures deployed by South Africa impose large economic costs and have negative implications for the factor distribution of income. Labor with low education levels are much more strongly affected than labor with secondary or tertiary education. As a result, households with low levels of educational attainment and high dependence on labor income would experience an enormous real income shock that would clearly jeopardize the food security of these households. However, in South Africa, total incomes for low income households are significantly insulated by government transfer payments. From public health, income distribution and food security perspectives, the remarkably rapid and severe shocks imposed because of Covid-19 illustrate the value of having in place transfer policies that support vulnerable households in the event of ‘black swan’ type shocks. |
| COVID-19 pandemic and mitigation strategies: implications for maternal and | Akseer, N., Kandru, G., Keats, E. C., & Bhutta, Z. A. | 2020-08 | The American Journal of | https://doi.org/10.1093/ajcn/nqaa171 | Coronavirus disease 2019 (COVID-19) continues to ravage health and economic metrics globally, including progress in maternal and child nutrition. Although there has been focus on rising rates of childhood wasting in the short term, maternal and child undernutrition rates are also likely to increase as a consequence of COVID-19 and its impacts on poverty, coverage of essential interventions, and access to appropriate nutritious foods. Key sectors at particular risk of collapse or reduced efficiency in the wake of COVID-19 include food systems, incomes, and social protection, health care services for women and children, and services and access to clean water and sanitation. This review highlights key areas of concern for maternal and child nutrition during and in the aftermath of COVID-19 while providing strategic guidance for countries in their efforts to reduce maternal and child undernutrition. Rooted in learnings from the exemplars in Global Health's Stunting Reduction Exemplars project, we provide a set of recommendations that |

| | | | | | |
|---|---|---|---|---|---|
| child health and nutrition. | | | Clinical Nutrition | | span investments in sectors that have sustained direct and indirect impact on nutrition. These include interventions to strengthen the food-supply chain and reducing food insecurity to assist those at immediate risk of food shortages. Other strategies could include targeted social safety net programs, payment deferrals, or tax breaks as well as suitable cash-support programs for the most vulnerable. Targeting the most marginalized households in rural populations and urban slums could be achieved through deploying community health workers and supporting women and community members. Community-led sanitation programs could be key to ensuring healthy household environments and reducing undernutrition. Additionally, several COVID-19 response measures such as contact tracing and self-isolation could also be exploited for nutrition protection. Global health and improvements in undernutrition will require governments, donors, and development partners to restrategize and reprioritize investments for the COVID-19 era, and will necessitate data-driven decision making, political will and commitment, and international unity. |
