## Supplementary File 5 - Selected Literature - Addressed Areas of Research and Study Designs for "The COVID-19 pandemic and child malnutrition in sub-Saharan Africa: A scoping review"

### Selected Papers - List of Addressed Areas of Research and types of publications

| | Type of Publication / Study Design | Funding Source | Main area of research and identified socioeconomic factors leading to a COVID-19 induced rise in malnutrition | Priority areas for action on malnutrition mitigation |
|---|---|---|---|---|
| Roberton T., et al., 2020 | Scenario Analysis | Bill & Melinda Gates Foundation, Global Affairs Canada. | • COVID-19 induced reduction in household income/unemployment leading to a rise in malnutrition rates<br>• Three scenarios of covid-19 effects - including reduction in coverage of essential maternal and child health interventions reduction by 9·8–51·9% and the prevalence of wasting is increased by 10–50% and induced rise in mortality.<br>• Increase in under-5 child mortality through diminished access to health services and access to food due to COVID-19 pandemic.<br>• Decrease in access to food through unavoidable shocks, health system collapse and/or intentional choices conducted in the context of the COVID-19 pandemic<br>• Disruption of routine health care and dimished access to food and resulting increase in child and maternal deaths. | • Formulation of informed pandemic-related policy decisions on the basis of statistical modelling, as COVID-19's direct effects on pregnant women and infants have already been estimated using models.<br>• There is currently insufficient accurate empirical evidence on the impact of the pandemic on the provision and utilization of health services.<br>• The authors stress that the findings are meant to aid in understanding the possible magnitude of the indirect effects of the pandemic, not to provide precise statistics or advise. |
| Jafri A., et al., 2020 | cross-sectional | No funding received | • Perceived effects of food availability, accessibility, and dietary behavior | • The first step should be to implement emergency livelihoods |

| | | | | |
|---|---|---|---|---|
| | multi-country online survey (quantitative) | | changes through the pandemic lockdown.<br>• Stockpiling of food<br>• Increase in the prices of staple foods resulting in difficulties in sufficient food acquisition<br>• The increase in prices is explained by the authors through domestic demand being stifled by increased transaction costs. Import declines can also result in higher prices and a shortage of basic consumer items, which can in the long-term also contribute to higher inflation.<br>• Diminished food variety, quality and quantity<br>• Vulnerable groups – children, elderly and people with chronic disease – reported to have vastly diminished rates of food acessibility | and food support interventions to ensure that the most vulnerable people have access to food. In times of disaster, emergency livelihoods is a type of livelihood intervention that adapts to meet the immediate needs of the most vulnerable communities.<br>• Emergency short-term employment/cash-for-work geared to COVID-19 preventive measures such as building and rehabilitating public sanitation, disinfecting and cleaning public buildings, providing food assistance, and financial grants for the most vulnerable households are examples of initiatives.<br>• COVID-19 has led to a vast amount of new vulnerable groups, including casual workers, small business owners, and employees in the private sector who have lost their employment and income. These new vulnerable populations should be included to the list of beneficiaries of food and social assistance programs. Changes in food costs have an impact on these new vulnerable groups, thus |

| | | | | |
|---|---|---|---|---|
| | | | | strategies to promote access to nutritious meals are needed.<br>• In addition to establishing price control measures that aim to minimize the burden on the most impacted, access to accurate food price information should be prioritized as a policy response. Governments should also improve food price monitoring and reinforce enforcement of food pricing laws that are broken.<br>• Food production, distribution, and marketing should be monitored using rapid and repeated assessment methodologies in order to provide real-time data for evaluating food security impacts and informing post-COVID-19 recovery and crisis management. |
| Govender K., et al., 2020 | Literature Review | No funding received | • The informal sector (The part of a nation’s economy whose activities are not recorded in official statistics) provides income to the majority of the region's employable young people. Stringent public health control measures associated to COVID-19 are worsening hunger and poverty among young people in countries that had food | • Governmental food subsidy programs for vulnerable groups living in poverty<br>• NGOs with established networks are more likely to have access to marginalized groups, and they should function as intermediaries between recipients and funders, providing shelter, food, and other critical services. |

| | | | | |
|---|---|---|---|---|
| | | | insecurity prior to the epidemic (e.g., Zimbabwe).<br>• The closure of non-governmental organizations (NGOs) and community centers puts even more strain on the homeless and street children who rely on them for food, clothing, and basic hygiene supplies. | • Food insecurity can be alleviated by addressing the impact of income loss in lower-income households by allocating financial transfers. South Africa, for example, has adopted the COVID-19 Social Relief of Distress grant, which is granted to unemployed people who do not receive any other type of social assistance.<br>• Food aid programs (such as food banks) should be widely implemented in both urban and rural regions to augment cash transfers, assure access to life-sustaining food, and prevent social unrest and hunger riots.<br>• Providing direct livelihood support to low-income households through sponsored programs to enhance small-scale livestock and agricultural operations will boost child nutrition while also reducing the impact of food shortages and price rises. |
| Egger D., et al., 2021 | Meta-Analysis | Energy for Economic Growth (EEG), UNOPS Sierra Leone, Bill & | • Impediments to livelihood have exacerbated the negative economic shock experienced by those polled in these nations. In most nations, a high percentage of respondents say they | • Because of the limited contact required to implement, recent inventions to swiftly and safely identify the impoverished using mobile phones or satellite data and |

| | | | | |
|---|---|---|---|---|
| | | Melinda Gates Foundation | have less access to markets, with the median share being 31% (range 3–77%) (column 3), which is likely due to the widespread lockdowns and other mobility restriction regulations implemented from March to June 2020.<br>• These decreases in employment, income, and market and service access all appear to be contributing to rising levels of food insecurity. Between 9 and 87 percent of respondents were compelled to skip or minimize meals throughout the study period. | send donations remotely through mobile money transfers have promise in this context.<br>• In the event that COVID-19 disease environment or the associated economic downturn persists for an extended period, policymakers in LMICs will need to devise inventive strategies to build income-generating enterprises with longer gestation periods.<br>• For example, "graduation programs" that combine assets and training can encourage a source of income with minimal external engagement and have already been demonstrated to lessen poverty. Combining these programs with immediate cash assistance has also been found to aid in the development of long-term income sources during times of civil upheaval. |
| Fore, H. et al., 2020 | Literature Review | No funding received | • Physical separation, school closures, trade restrictions, and country lockdowns are some of the COVID-19 response strategies that are affecting food systems by disrupting the production, transportation, and sale of nutritious, fresh, and affordable foods, | • Protect and promote access to healthy, cheap, and nutritious foods.<br>• Invest in improving maternal and child nutrition during pregnancy, infancy, and early childhood |

| | | | | |
|---|---|---|---|---|
| | | | forcing millions of families to rely on nutrient-poor alternatives.<br>• Access to crucial and often life-saving nutrition services is deteriorating due to overburdened health systems and pauses in humanitarian response. In many LMICs, social protection systems are overburdened as families in need struggle to get the food and services they require during economic downturns.<br>• COVID-19's effects in mothers and children may be exacerbated by malnutrition. At the same time, more children are becoming malnourished as the quality of their diets deteriorates, as well as disruptions in nutrition and other important services, as well as the socioeconomic shocks caused by the epidemic in LMICs.<br>• According to new estimates published in The Lancet by Derek Headey and colleagues, if no action is taken soon, the global frequency of child wasting might grow by a startling 143%. Before the COVID-19 epidemic, an estimated 47 million children under the age of five were affected by wasting. During the first 12 months of the epidemic, an estimated 67% of children in Sub- | • Re-activate and scale up services for the early detection and treatment of child wasting<br>• Maintain the provision of nutritious and safe school meals for vulnerable children<br>• Increase social protection to ensure that people have access to healthy foods and needed services. |

| | | | | |
|---|---|---|---|---|
| | | | Saharan Africa and South Asia will be wasting, with 80% of them in Sub-Saharan Africa and South Asia. | |
| Headey D., et al., 2020 | Literature Review | Children's Investment Fund Foundation (CIFF) | • Due to severe drops in household incomes, changes in the availability and price of nutritious foods, and pauses in health, nutrition, and social protection services, a rise in child malnutrition, particularly wasting, is projected.<br>• Even relatively short lockdown measures, when paired with severe mobility disruptions and comparatively mild food system disruptions, result in a 79% (SD 24%) drop in GNI per capita in most LMICs compared to pre-COVID-19 predictions.<br>• Estimates from microeconomic models show that drops in GNI per capita are linked to drastical increases in child wasting.<br>• COVID-19-related estimated country-specific losses in GNI per capita might result in a 143% rise in the prevalence of moderate or severe wasting among children younger than 5 years, according to estimations applied to 118 LMICs. In 2020, we expect that there will be an additional 67 million children with wasting, compared to forecasts for | • At a time when most country economies are hurting from COVID-19-related losses, a quick mobilization of domestic and donor resources is required. The World Bank estimated in 2017 that achieving the global Sustainable Development Goal nutrition targets will require $7 billion per year for the next ten years. To overcome COVID-19-related difficulties, these projections must be increased upwards. |

| | | | | |
|---|---|---|---|---|
| | | | 2020 without COVID-19, an estimated 21·8% in sub-Saharan Africa.<br>• When the projected increase in wasting in each country is combined with a projected year-average reduction of 25% in nutrition and health-care coverage, we estimate that there will be 128 605 additional deaths in children younger than 5 years in 2020 (ranging from 111 193 to 178 510 for best and worst case scenarios), with 52 percent of these deaths occurring in Sub-Saharan Africa. | |
| Coker M. et al., 2020 | Literature Review | No funding received | • Lockdowns combined with school closures have hampered access to school-based meals, which are one of the few reliable sources of food for many children. As a result of the epidemic, children have been exposed to malnutrition, poor nutrition, and, as a result, harmful effects on cognitive development; all of this comes at a time when many families are facing unemployment and income loss.<br>• According to the World Food Programme, 368 million children aged pre-primary to secondary school (47 percent girls) worldwide are currently missing school meals, with 148 million in Sub-Saharan Africa. In SSA, where | • Strengthening national food, health, and social protection programs is encouraged by the WHO's Action Plan on Child Wasting and the African Union's Nutrition Strategy. However, in this emergency pandemic scenario, palliatives of food and financial assistance must be delivered as soon as possible , with the most vulnerable children and families receiving priority.<br>• In SSA nations, public health responses to the epidemic are growing; nonetheless, attention to socioeconomic determinants of health is woefully lacking. |

| | | | | |
|---|---|---|---|---|
| | | | school meals are typically a substantial incentive for parents to enroll female children and so prevent early child marriage, the effects may be much greater. | Governments should implement measures such as improved access to courts, legal protection, and housing to address the needs of vulnerable children as an ethical obligation. No. 112 |
| Zar H. et al., 2020 | Literature Review | South Africa Medical Research Council | • Children in LMICs are not at high risk for severe COVID disease, as they are in HICs, but there are significant detrimental indirect impacts on child health.<br>• One of COVID's indirect consequences on child health in LMICs is the downscaling or closure of routine child and maternal health preventive and other services, which jeopardizes nutritional programs and leads to increased morbidity and death.<br>• Many children from underprivileged neighborhoods in LMICs rely on school food programmes for daily nourishment, putting their nutrition at risk.<br>• Increased poverty, disrupted schooling, lack of access to school food systems, restricted access to health facilities, and disruptions in vaccination and other child health initiatives are all indirect effects of the epidemic that are cause for concern. | - |

| | | | | |
|---|---|---|---|---|
| | | | • lockdowns have a significant economic impact, resulting in widespread unemployment, food insecurity, and housing insecurity, further jeopardizing health and escalating intrafamilial violence. | |
| Aborode A. T., et al., 2020 | Literature Review | No funding received | • According to the International Food Policy Research Institute, an additional 140 million people will be forced into extreme poverty in 2020 as a result of the pandemic, living on less than $1.90 per day. According to the World Food Programme, by the end of 2020, the number of people in LMICs experiencing acute food insecurity will have nearly doubled to 265 million.<br>• Because of the restraints in mobility and transportation, widespread hunger and malnutrition are likely to worsen. COVID-19 is spreading across Africa amid alarming levels of famine and malnutrition, which are likely to intensify as the virus threatens livelihoods and family economics.<br>• In these areas, the young children aged 1 to 5 years are the ones who are most affected by demography. Physical separation, school closures, trade restrictions, and country lockdowns | • To safeguard these children, prevent and treat hunger, and reduce human loss, UNICEF estimates that a minimum of US$2.4 billion is required immediately. A total of $2.4 billion is estimated to be spent on a package of four life-saving interventions:<br> ○ prevention of wasting in at-risk children. When schools are closed, continue to provide healthy and safe school meals for disadvantaged children through home delivery, take-home rations, and cash or vouchers. These efforts must also guarantee that school meals or food packages have enough nutritional value and that harmful foods and beverages are not provided. |

| | | | | |
|---|---|---|---|---|
| | | | have all been implemented as ways to combat the pandemic.<br>• They have, however, exacerbated food insecurity by reducing the production and sale of nutritious and inexpensive food products, forcing millions of people to rely on nutrient-poor alternatives. Access to crucial and often life-saving nutrition services is deteriorating due to overburdened health systems and pauses in humanitarian response.<br>• Millions of children are at risk of not receiving the care they need to survive and thrive because services for the prevention and treatment of wasting have been largely disrupted in LMICs. According to UNICEF data from the early months of the COVID-19 pandemic, coverage of critical nutrition services in LMICs was reduced by 30%, with falls of 75–100% in lockdown situations.<br>• With an estimated 47 million children under the age of five years affected by wasting globally prior to the COVID-19 pandemic, this would imply an additional 6.7 million children with wasting during the first 12 months of the pandemic, 80% of whom would be | ○ As a cornerstone of the COVID-19 response, access to nutritious, safe, and affordable meals must be protected and promoted. Protecting food farmers, processors, and retailers; discouraging trade bans;<br>○ treatment of wasted youngsters<br>○ biennial vitamin A supplementation for children aged 6–59 months (90 percent coverage)<br>○ mass communication for the protection, promotion, and support of breastfeeding that focuses on caregivers or families of children aged 0–23 months. In the context of COVID-19, protect breastfeeding and prevent the inappropriate marketing of infant formula to promote mother and child nutrition during pregnancy, infancy, and early childhood by enabling caregivers access to correct |

| | | | | |
|---|---|---|---|---|
| | | | in Sub-Saharan Africa and South Asia, and more than 10,000 additional child deaths per month during this time.<br>• Aside from an increase in marasmus, the COVID-19 pandemic is expected to result in an increase in other types of child malnutrition, such as stunting, nutritional deficiencies, and obesity. | information on newborn feeding |
| Nrambara J. & Chu M., 2021 | Descriptive Literature Review | No funding received | • Healthcare systems have struggled to maintain ordinary services in previous pandemics, according to the WHO, as people's efforts and medical supplies shift to respond to the emergency. As a result, basic and regular necessary health services, such as child nutrition, family planning, and food supplements to those in need, are frequently neglected, resulting in direct and indirect consequences on maternal and child nutrition.<br>• School closures have contributed to a disruption in their nutritional health because many children formerly got breakfast and lunch at school, which they were unable or challenged to obtain during COVID-19.<br>• Children spend far more time in front of television and computer screens, are more exposed to junk food and sugary beverage ads, consume junk food and | • One strategic response considers Increasing access to community-based nutrition initiatives that aid in the early detection and treatment of malnourished children, as well as emergency food delivery to poor families, especially those with children under the age of five, providing enriched vitamins and minerals.<br>• Community nutrition education in areas such as gestation, exclusive breast-feeding, and complementary feeding, as well as sanitary behaviors such as handwashing, adequate sanitation, and other basic behavioural adjustments, should be strengthened through counseling and promotion programs.<br>• Malnutrition can be controlled and eradicated with a multi-sectoral |

| | | | | |
|---|---|---|---|---|
| | | | sugary beverages, and participate in far less physical activity. | strategic approach. A multisectoral approach to avoiding childhood malnutrition will necessitate not only financial commitment, but also coordinated activities from several government ministries, UN-affiliated agencies, and non-governmental organizations. |
| Otekunrin A. et al., 2020 | Literature Review | No funding received | • Despite the fact that there are fewer occurrences of COVID-19 among children with weaker symptoms, the country's 156 actions to the illness outbreak can have serious effects for child nutrition and educational 157 results.<br>• Due to declining income, high levels of malnutrition among women, children, and the elderly would most likely worsen in poor families across Africa.<br>• People in food crises will have their health and nutrition threatened even more due to their difficulty to access already overburdened health care and their inability to move around to fend for themselves.<br>• Children whose families rely on the government's home-grown school feeding program to supplement their children's meals would experience | • In the face of the COVID-19 epidemic, African countries should prioritize sustainable agricultural practices while paying close attention to the creation and execution of policies that minimize hunger.<br>• In this phase of COVID-19, there is still a shortage of focus on sustainable agriculture, which can enhance nations' agricultural productivity. (supply chain enhancement)<br>• Farmers of small scale must be allowed to continue their businesses (South Africa, Nigeria) Farming activities, in order to reduce agricultural productivity decline and allow for a stabilization of Food production, distribution, and supply during the rest of the pandemic. |

| | | | | |
|---|---|---|---|---|
| | | | more malnutrition as a result of movement limitations.<br>• If food supply networks are not adequately coordinated, they may suffer negative consequences. Funds intended to improve the agriculture sector are likely to be transferred to help battle COVID-19. | • Informal food traders, grocery stores, wholesale produce markets, and food markets must continue to operate in order to provide adequate food supply in the proper amount and quality.<br>• Temporary food markets are set up in neighborhoods to improve physical access to food.<br>• Payments of social subsidies and financial transfers to vulnerable households<br>• To help firms and employees cope with the impact of COVID-19 on their finances, tax measures such as tax subsidies, employment tax incentives, debt relief finance schemes, loan repayment exemptions, trade policies, and so on have been introduced (South Africa, Nigeria) |
| Kansiime M. K. et al., 2021 | Quantitative Online Survey | CABI Development Fund (CDF) (supported by contributions from the Australian Centre for International | • household income and food security in two East African nations – Kenya and Uganda – were assessed, as a result of the Coronavirus Disease 19 (COVID-19) pandemic (442 participants)<br>• The COVID-19 crisis caused income shocks for more than two-thirds of the respondents. Food insecurity and dietary quality have deteriorated. | • Government responses should concentrate on structural improvements in social security, such as designing responsive packages to help people who have been forced into poverty as a result of pandemics.<br>• Building financial institutions to aid in the medium-term recovery of |

| | | | | |
|---|---|---|---|---|
| | | Agricultural Research, UK's Foreign, Commonwealth & Development Office (FCDO), the Swiss Agency for Development and Cooperation and others | • Because for some kids living in poverty, schools are not just a place to learn but also a place to eat properly, having children at home due to school closures is more likely to worsen food insecurity.<br>• In Kenya and Uganda, the proportion of people who are food insecure has climbed by 38% and 44%, respectively.<br>• In comparison to other respondent categories, income-poor households and those relying on labor income were more vulnerable to income shock and had inferior food consumption during the COVID-19 pandemic.<br>• Higher food prices, along with increased consumption and high reliance levels, will undoubtedly have greater socio-economic consequences, particularly for the most disadvantaged households. | enterprises, as well as guaranteeing the resilience of food supply chains, particularly those that provide nutrient-dense foods.<br>• Th e Government of Kenya (GoK) announced a 100% tax relief for individuals earning a gross monthly income of KES 24,000 (USD 230) or less, a reduction of the income tax rate (Pay As You Earn - PAYE) from 30% to 25%, a reduction of resident income tax (Corporation Tax) from 30% to 25%, and a reduction of the turnover tax rate from 3% to 1% for all small businesses (SMEs) , as well as the removal of loan defaulters from the Credit Reference Bureau's (CRB) database. |
| Arndt C., et al., 2020 | Quantitative social accounting matrix | No funding received | • South Africa's distancing measures come at a high cost to the economy and have a detrimental impact on the income factor distribution. Low-skilled workers are disproportionately affected compared to those with a secondary or university degree. As a result, households with low levels of educational attainment and a large reliance on work income would face a | • The extremely swift and severe shocks inflicted by Covid-19 demonstrate the need of putting in place transfer programs that help poor households from the viewpoints of public health, income distribution, and food security.<br>• In South Africa, government transfer payments protect low- |

| | | | | |
|---|---|---|---|---|
| | | | significant real income shock, putting their food security in jeopardy.<br>• This first effect on household income occurs when extremely minor direct affects on food production, no influence on food pricing, and no influence on food distribution networks are assumed.<br>• The lowest two education levels have a 40% drop in pay earnings, which translates to a 40% drop in hours worked. These losses are definitely substantial to endanger food security for a home depending on wage earnings from low-educated workers, especially if the household was previously disadvantaged. As you move up the income scale, the impact on hours worked for tertiary educated workers is less severe (26%) since companies that rely on on highly trained workers may adjust to lockdown with more flexible work arrangements. | income households' overall incomes greatly.<br>• |
| Nechifor V. et al., 2021 | | No funding received | • The economic consequences of the COVID-19 quarantine could undo recent gains in food security. The lockdown in April–June may have resulted in a significant drop in GDP and household | • Food and financial assistance programs for the most vulnerable people.<br>• Kenya's fiscal and public spending policies have helped to mitigate some of the negative welfare |

| | | | | |
|---|---|---|---|---|
| | | | income, resulting in diminished demand for food items.<br>• The negative impact of decreasing exports on the trade balance and the domestic exchange rate affects the affordability of imported food commodities, resulting in decreased consumption of staple food products. | effects that have a direct influence on food security. However, the data suggest that calorie intake and macronutrient balance increase unevenly among household groups, with rural households and those with child stunting showing the least change.<br>• Scaling up the cash payments program and expanding the beneficiary pool to the poorest income percentiles is one important route for intervention to enhance short-term food security outcomes. This represents the majority of the people residing in rural areas, who had low calorie intakes prior to the pandemic and whose income and food consumption have yet to recover. |
| Akseer N., et al., 2020 | Narrative Literature Review | Gates Ventures | • Food systems, incomes, and social protection, health care services for women and children, and services and access to clean water and sanitation are among the key areas at danger of collapse or reduced efficiency as a result of COVID-19.<br>• Furthermore, with restricted access to fresh produce, children and families may be more inclined to turn to | • While long-term agrarian land reforms focused on transferring land ownership and adopting innovative/efficient agriculture techniques may reduce undernutrition, urgent interventions are equally important. One of Ethiopia's food-insecurity solutions (the Productive Safety Net Program) was designed |

| | | | | |
|---|---|---|---|---|
| | | | cheaper and more readily available processed and prepackaged, high-sodium, and less-nutritious meals, which can have negative health repercussions.<br>• Low agricultural production and disruptions in the food import–export system disrupt local food markets and small companies, while loss of family income exposes vulnerable households to price spikes and food shortages.<br>• Despite being ostensibly "free" from lockdowns, COVID can have direct and indirect effects on the functioning of the food supply chain in LMICs, particularly in the informal sector.<br>• While immediate effects, such as restaurant closures and vendor limitations, account for a tiny portion of the entire food economy in urban areas, the impact on rural markets might be much bigger.<br>• People in LMICs have been hit hard by indirect effects such as unemployment and decreased salaries of daily wage laborers and industry employees.<br>• Access to routine health treatments for women and children has deteriorated greatly as a result of overcrowded health systems, restricted travel, and | to provide emergency food help to 15 million people who were at risk of hunger, and it was regarded crucial to the country's stunting-reduction story. In COVID-19-affected nations, such long- and short-term solutions addressing both supply and demand-side concerns might be considered for nutrition protection.<br>• An effective community health extension system may provide access to health care for even the most rural and hard-to-reach communities, as has been demonstrated in numerous stunting-reduction model nations. Ethiopian health extension workers (HEWs) and Nepalese female community health volunteers (FCHVs) exemplify how community health workers (CHWs) may offer immunizations, nutritional supplements, health and nutrition education, and even reproductive, maternal, and newborn care. Rather than relying on pure volunteering, it is currently recommended that such CHWs be paid. While the primary health care |

| | | | | |
|---|---|---|---|---|
| | | | shifting priorities at the primary care level.<br>• Prior to COVID-19, quality of care was a continuous issue; nevertheless, in its present condition and for years to come, focused initiatives for high-quality health care for those in the most need will likely take a backseat. As a result, the health and danger of undernutrition in women and their children may substantially worsen, particularly if existing conditions persist for an extended period of time. | system may not be entirely functioning and resources are few during the COVID-19 crisis, governments may explore repurposing current CHW cadres to address increasing maternity, child health, and nutrition screening in communities. These CHWs are also crucial in re-establishing community-based malnutrition control initiatives.<br>• The CHW program in Senegal (24) has shown to be a successful means of disseminating health best practices to the general public. The FCHV (19) and HEW (18) programs in Nepal and Ethiopia, respectively, have had very successful health and nutrition counseling components. |
